# The impact of HIV on women living with HIV and their families in low- and middle-income countries: A systematic review

**DOI:** 10.1101/2022.04.16.22273930

**Authors:** Nelsensius Klau Fauk, Lillian Mwanri, Karen Hawke, Leila Mohammadi, Paul Russell Ward

**Affiliations:** Torrens University Australia, Adelaide, 88 Wakefield St, Adelaide SA 5000, Australia; Institute of Resource Governance and Social Change, Jl. R. W. Monginsidi II, No. 2, Kupang, Indonesia, 85221; Aboriginal Communities and Families Research Alliance, South Australian Health and Medical Research Institute, Australia; College of Medicine and Public Health, Flinders University, GPO Box 2100, Adelaide 5001, Australia

**Keywords:** WLHIV, HIV impact, HIV-affected families, low- and middle-income countries

## Abstract

HIV infection adds a significant burden to women in low- and middle-income countries (LMICs), often leading to severe detrimental impact, not only on themselves, but also on their families and communities. Given that more than half of all people living with HIV globally are females (53%), this review seeks to understand the impact of HIV infection on women living with HIV (WLHIV) and their families in LMICs, and the interrelationships between one impact and another. A systematic review was conducted to find literature using the following databases: Medline, PsycINFO, CINAL, Emcare, Scopus and ProQuest. Research articles were included if they met the following inclusion criteria: conducted in LMICs, published in English language between January 1^st^ 1990 and October 31^st^ 2021, had full text available, involved WLHIV (married and unmarried), and focused on the impact of HIV on these women and their families. Critical appraisal tools developed by Joanna Briggs Institute (JBI) were used to assess the methodological quality of the studies and thematic narrative synthesis was used to analyse the findings. A total of 22 articles met the inclusion criteria. The review showed that HIV has a range of negative consequences on WLHIV and their families including: (i) psychological impact, (ii) poor physical health and intimate partner violence, (iii) social impact, and (iv) economic impact. The findings indicate the need for targeted interventions, specific to WLHIV, that address the inequity and discrimination they face. These interventions should also incorporate education and sustainable support structures for WLHIV and their families.

## 1. Introduction

The Human Immunodeficiency Virus (HIV) and the Acquired Immune Deficiency Syndrome (AIDS) have been a worldwide public health problem for close to four decades (1). The 2020 UNAIDS report shows an estimated 37.7 million people living with HIV (PLHIV) worldwide, 1.7 million new diagnoses and 680,000 AIDS-related deaths in 2020 (1, 2). Globally, 53% of the estimated number of PLHIV are women aged 15 and over (2). In sub-Saharan Africa (SSA) where the majority (70.5%) of PLHIV reside, women living with HIV (WLHIV) represented 63% of HIV infections among adults aged 15 and older (1, 2). The current report also shows an estimated 4200 women aged 15-24 years becoming infected with HIV every week and are twice likely to be living with HIV than men in SSA (2). In 2020 in Asia and the Pacific region, WLHIV aged 15-24 years and 25-49 years represented 11% and 19%, respectively, of the total number of 5.8 million PLHIV (1). Globally, around 5000 women aged 15-24 years become infected with HIV every week (1).

It is known that compared to men or other key-affected populations such as transgender populations and men who have sex with men in low- and middle-income countries (LMICs), women generally face a greater burden of HIV impact (3–9). LMICs are the countries defined by the World Bank as those with gross national income per capita between $1,045 or less (low-income), $1,045 and $4,095 (lower middle-come) and $4,096 and $12,695 (upper middle-income) (10). Previous studies have reported that WLHIV in LMICs experience considerable psychological challenges, including depression, stress, anxiety and fear due to various concerns facing them following the HIV diagnosis (1, 11, 12). WLHIV are also reported to experience a number of social challenges such as stigma and discrimination by others or non-infected people within families, communities, workplaces and healthcare settings, manifesting in various discriminatory and stigmatising attitudes and behaviours (7, 11, 13). A lack of knowledge about how HIV is transmitted and prevented and the fear of contracting HIV through physical, social and healthcare-related contacts have often been reported as the main supporting factors for such stigma and discrimination against WLHIV (14, 15). For example, these lead to WLHIV being negatively labelled, avoided, rejected and excluded by others within families, communities and healthcare settings due to their HIV-positive status (16–18). Receiving a diagnosis and living with HIV also have negative economic consequences on women and their family through several mechanisms, such as women’s inability to work due to poor physical health condition, loss of employment leading a reduction in or loss of family incomes, and increased health expenditure (9, 19, 20). Such economic consequences have been reported to also lead to food insecurity within the family of WLHIV, forced sale of family properties including land and houses to cover healthcare and living expenses (9, 19, 21–23). HIV diagnosis among women or mothers also causes negative outcomes for education and wellbeing of their children (19, 24, 25), and child-mother separation or child-parent conflict which can have an extremely detrimental effect on the children’s mental health (20, 25).

Despite previous studies reporting how HIV affects the lives of PLHIV, to date, there have been no published systematic reviews, which examine the impact HIV has on WLHIV and their families in LMICs. This study aims to improve our understanding of HIV impact on WLHIV and their families, the mechanisms through which HIV impact influences their lives and interrelationships between one impact and another. It is also crucial to gain greater understanding of the consequences of an HIV diagnosis for women and their families, and identify the drivers behind these consequences given the high burden of disease among WLHIV. Understanding and identifying HIV impact will support further research and advocacy around education, targeted evidence-based interventions, and healthcare systems that address the specific needs of WLHIV and their families.

## 2. Methods

### 2.1. The systematic search of literature

We performed an initial search of relevant key terms guided by the PICO (Population, Intervention, Comparison and Outcomes), a framework that has been used to inform evidence based practice (26). The main search was then drafted and refined in Medline using both Medical Subject Headings (MeSH) terms and keywords as described in the inclusion criteria section below. We then translated the search into multiple databases, including Medline, PsycINFO, CINAL, Emcare, Scopus and ProQuest.

### 2.2. Inclusion criteria

Studies were included if they: (i) were published in English and between Jan 1, 1990 and Oct 31, 2021 inclusive to capture evidence on the impact of HIV on women which seemed to emerge in 1990s (Appendix 1); (ii) were conducted in LMICs (based on the classification made by the World Bank); (iii) involved WLHIV (married and unmarried); and (iv) aimed to identify psycho-social, physical, health, economic and educational impact of HIV on WLHIV and their families (Figure 1). The keywords were used in combination using Boolean operator system including AND and OR. An example of full electronic search strategy in SCOPUS in presented below and the full key words or search strategy in all databases can be found in Appendix 1.

**Figure 1.**
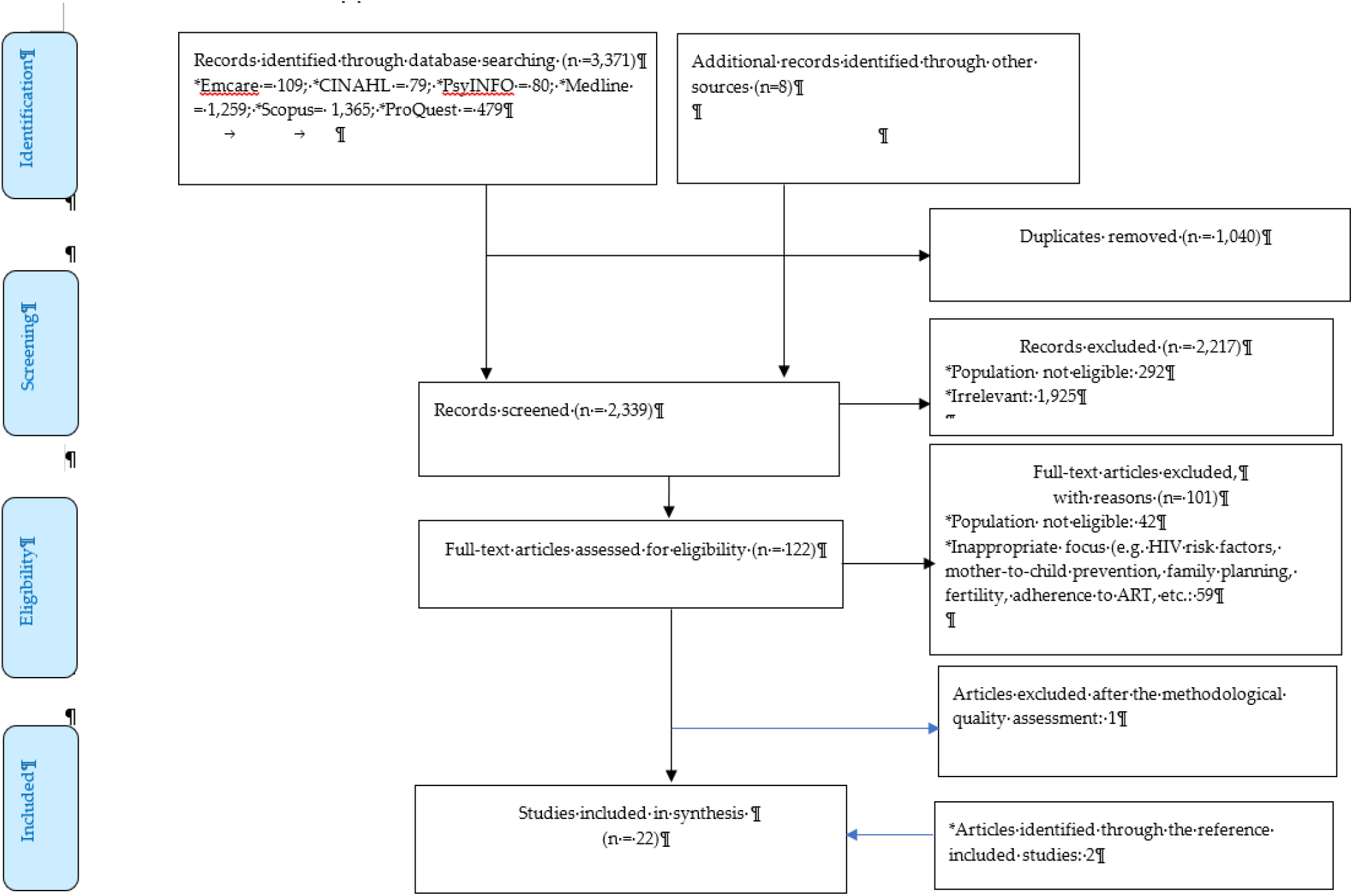
PRISMA Flow diagram of systematic literature search: records identified, screened, eligible and included in the review.

> TITLE-ABS-KEY((HIV* or “Human immunodeficiency virus” or AIDS)) AND TITLE-ABS-KEY((Wives or Wife or Mothers or female* or girl* or women or woman)) AND TITLE-ABS-KEY((predictor* or “risk factor*” or determinant* or “sexual behaviour” or “multiple sex partner*” or extramarital* or “sell* sex*” or “transactional sex” or prostitut* or “sex work” or condom* or “unsafe sex” or “unprotected sex” or knowledge or “social influenc*” or “peer influenc*” or “social norm*” or cultur* or sociocultural* or socioeconomic* or “social environmental*” or socioenvironment* or stigma or discriminat* or “psychological impact” or “social impact” or education or “school attendance” or “withdraw* from school” or stress or distress or depression or “psychosocial impact” or employment or “loss of job” or income or “nutrition security” or “food insecurity” or health or “physical health” or wellbeing or “healthcare accessibility*” or absenteeism or religio* or consequence*)) AND TITLE-ABS-KEY((family* or families)) AND TITLE-ABS-KEY(((Developing or “Less developed” or “low resource*” or disadvantaged or “resource limited” or poor or “low* or middle income*”) W/0 (countr* or region* or nation? or area*)))

### 2.3. Selection of the studies and methodological quality assessment

3,371 articles retrieved from the databases and eight from google through manual search were collated and imported into Endnote software (27). After removing 1,040 duplicates, 2,339 titles and abstracts were screened by two assessors (NKF and LM), further removing 2,217 articles not meeting the inclusion criteria. The assessment of full texts of remaining 122 articles led to a further removal of 101 articles not meeting including criteria and one article not meeting methodological quality. The reference listing of the 20 articles was scrutinised and two additional articles were obtained. The full texts of these two articles were also assessed to determine their eligibility. Twenty-two articles fulfilling the inclusion criteria and the methodological quality, were finally included in this review (Figure 1). The assessment for methodological quality was performed using critical appraisal tools developed by Joanna Briggs Institute (JBI) for study design (28). The methodological quality assessment was performed by two assessors (NKF and LM), and any disagreement between them was resolved through discussion. The appraisal forms for qualitative, cohort, cross sectional and case report studies comprised ten, nine, eight and eight questions respectively (Appendix 2). The questions were about the quality of the studies, for which each question received a response of Yes, No, Unclear, and Not Applicable.

### 2.4. Data extraction and analysis

Guided by the Thomas and Harden’s framework (29), a thematic analysis of selected articles was systematically performed as follows: (i) conducting a line by line open coding, with one assessor (NKF) extracting free codes from the findings of each article, (ii) developing descriptive themes where the free codes with similarity were organised or grouped together, (iii) reviewing (second assessor LM) of the initial descriptive themes, and thoroughly discussing (by both assessors – NKF and LM) any discrepancies, and (iv) finalising the review of descriptive themes and subthemes, with both assessors deciding the final analytical themes for the current study (29). The final version was agreed by all authors following further refinement of the theme and subtheme headings.

## 3. Results

### 3.1. Description of the included studies

Table 1 summarises the descriptive characteristics of the 22 included articles. Of the 22 articles, 12 studies (13 articles) were conducted in Asia and the Pacific region (6–9, 11, 16, 20, 30–35), six in Africa (5, 12, 13, 36–38) and one study (two articles) in the Caribbean (19, 25). One study (39) collected data from participants in Zimbabwe and the USA and presented the results of these two groups separately. For the purpose of this review, only the study results about the participants in Zimbabwe were included, as this fits the inclusion criteria. Of the 22 articles, thirteen were qualitative studies collecting data through one-on-one interviews (5, 6, 9, 12, 16, 19, 25, 31–35, 37). Nine articles used quantitative methods (7, 8, 11, 13, 20, 30, 36, 38, 39), of which seven were cross-sectional (7, 8, 11, 13, 36, 38, 39) and two were prospective cohort studies (20, 30). A total of 2,444 WLHIV participated in these studies, of whom 190 and 2,254 respectively were involved in qualitative and quantitative studies. Several studies involved a combination of WLHIV and children (25, 39), HIV negative women or women with unknown HIV status (13) and HIV positive men (8, 33), however only the results about WLHIV or the views of WLHIV were included in this review (Table 1). The sample sizes or the number of WLHIV who participated in the qualitative and quantitative studies reviewed varied from 1 to 30, and 50 to 633 people, respectively. The participants were recruited using convenient, purposive or snowball sampling techniques. Eight studies involved only married women (7, 9, 11–13, 20, 30, 34), nine involved a combination of married, widowed and single women (6, 8, 16, 31, 32, 36–39) and one study involved single women (33). Four studies did not report marital status of the participants (5, 19, 25, 35). Participants’ age ranged from 15 to 58 years old, however, three studies did not report participants’ age (8, 19, 25). The qualitative studies used content analysis (5, 12, 31, 32, 35) and thematic analysis (6, 16, 19, 25, 33, 34, 37) approach for data analysis, however one study employed a combination of thematic analysis, analysis of episode and identification of paradigm cases (9). The majority of the quantitative studies used one or a combination of bivariate and multivariate linear or logistic regression models for data analysis (7, 11, 13, 30, 36, 38, 39).

**Table 1.**
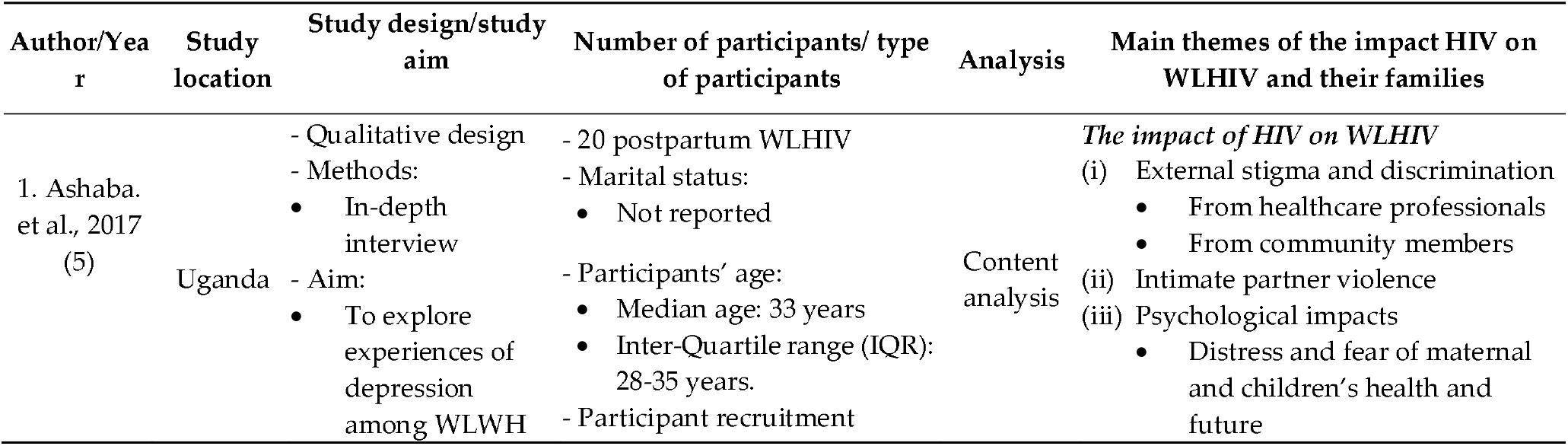
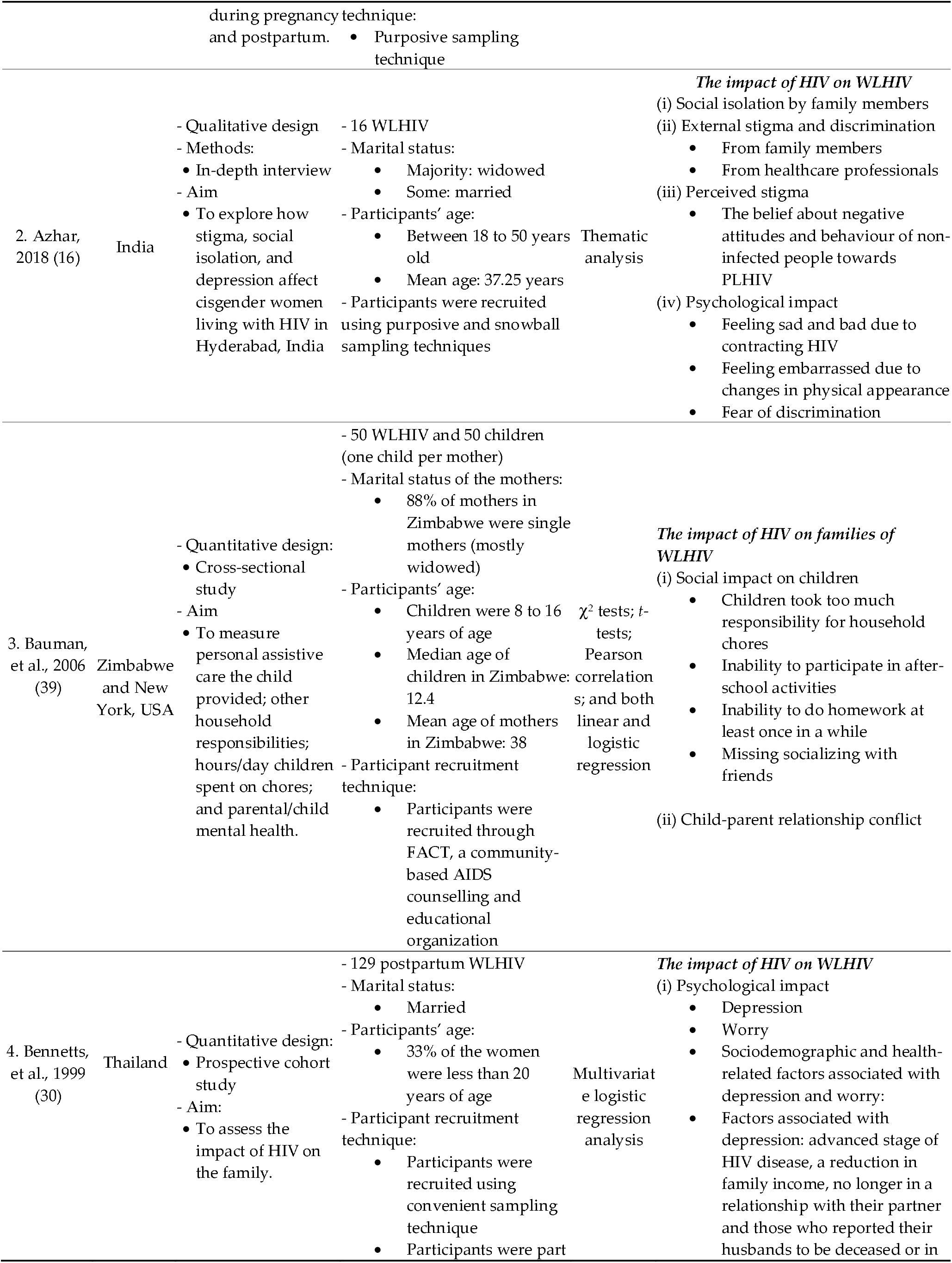
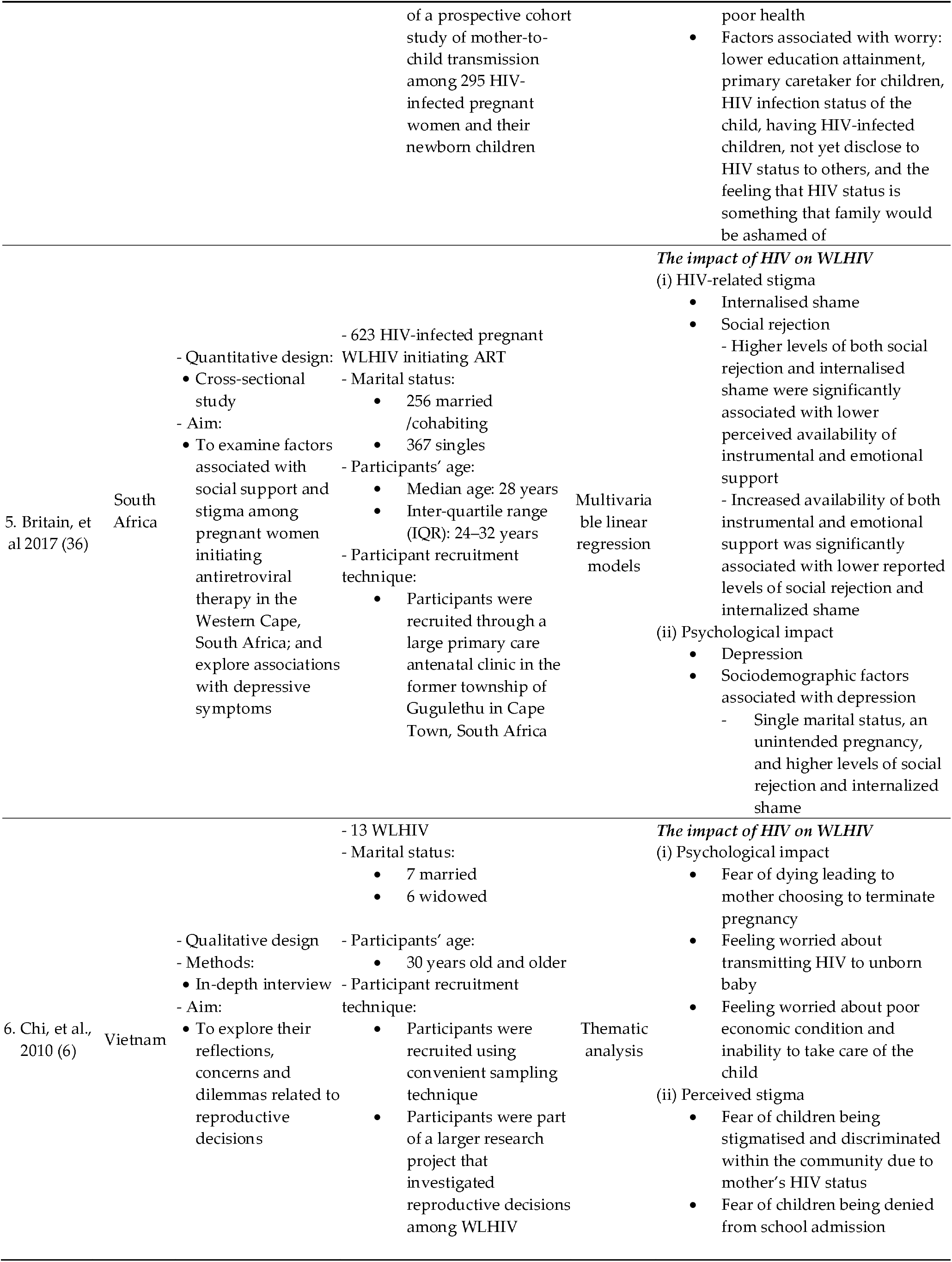
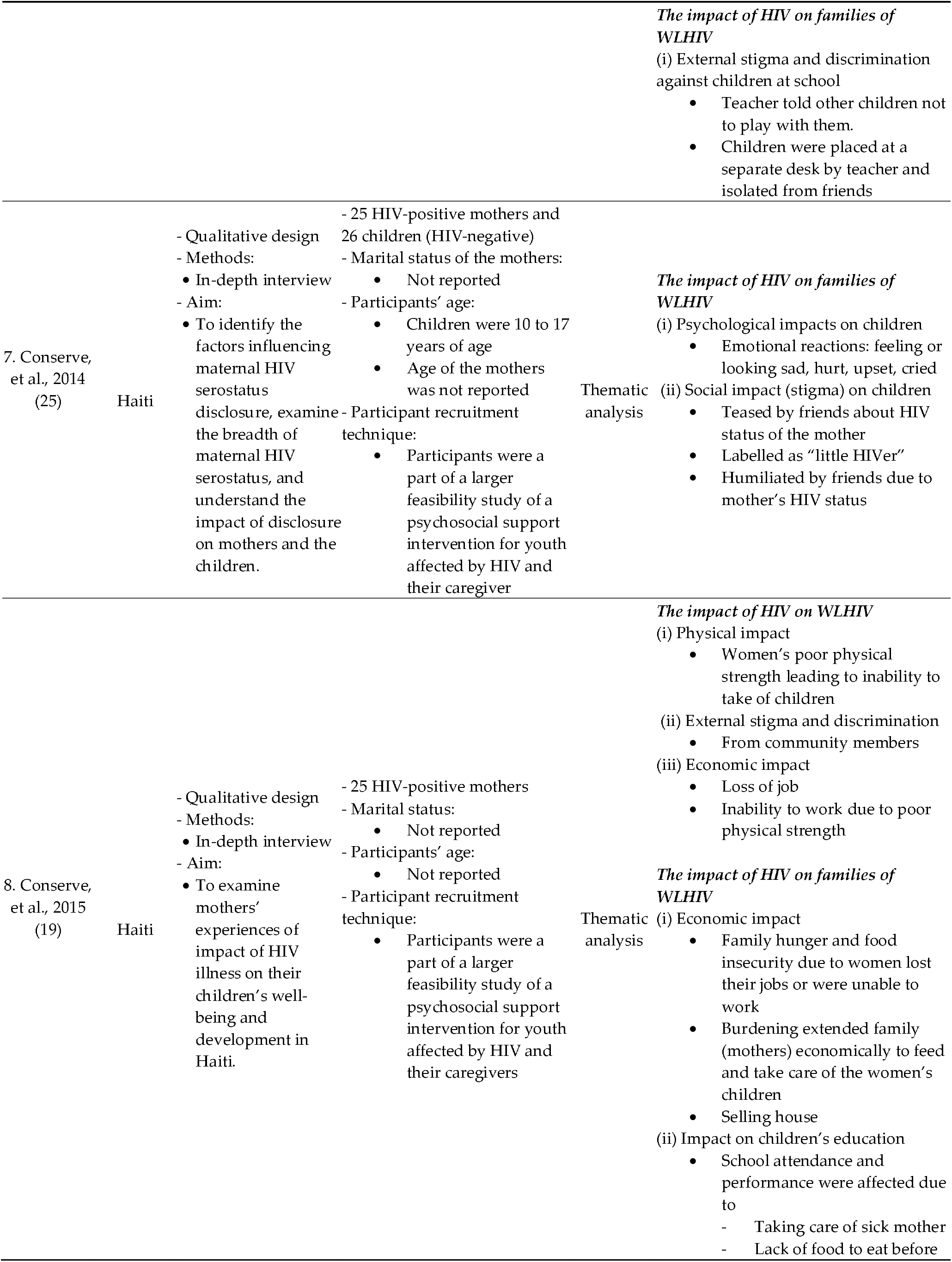
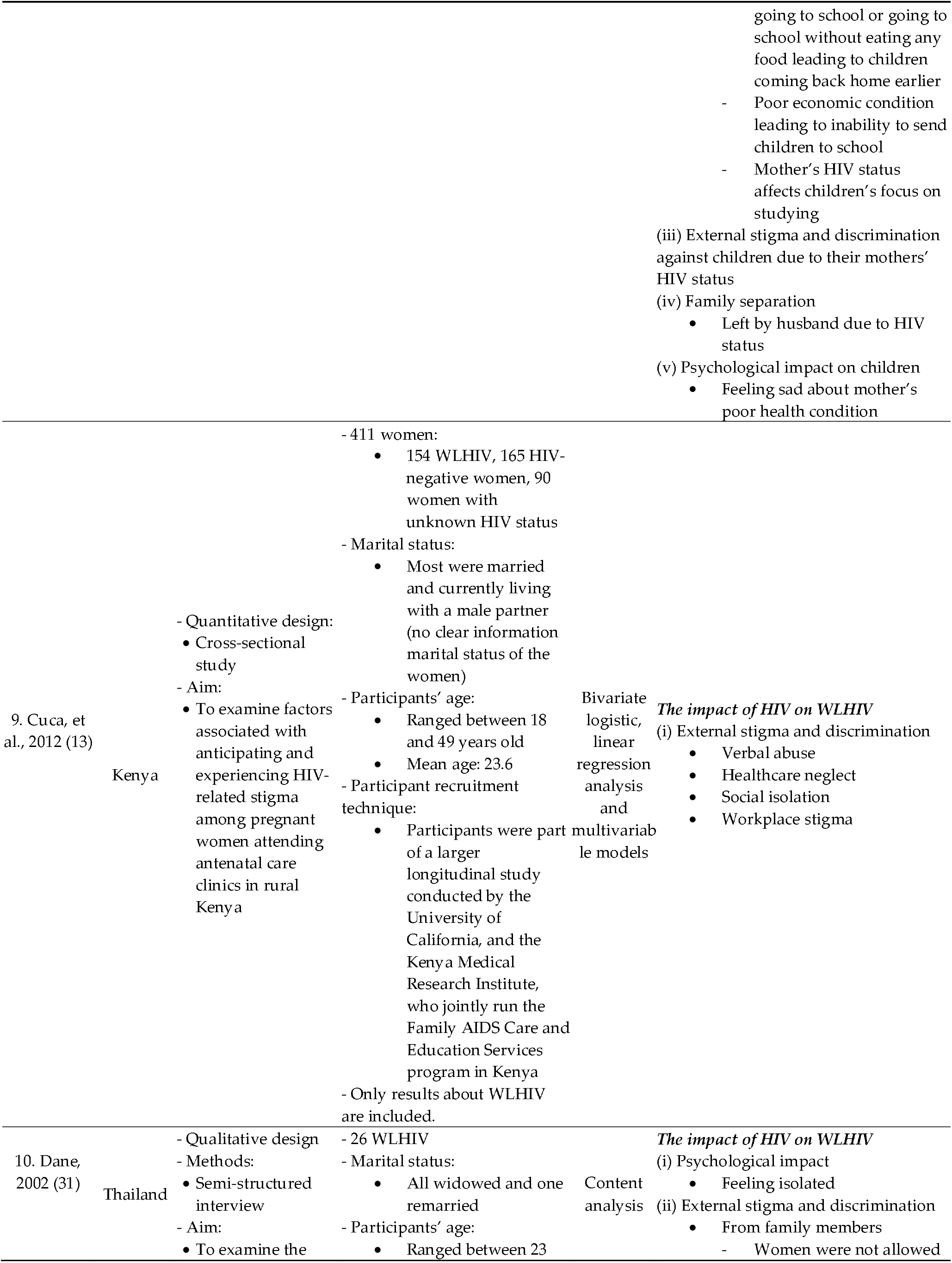
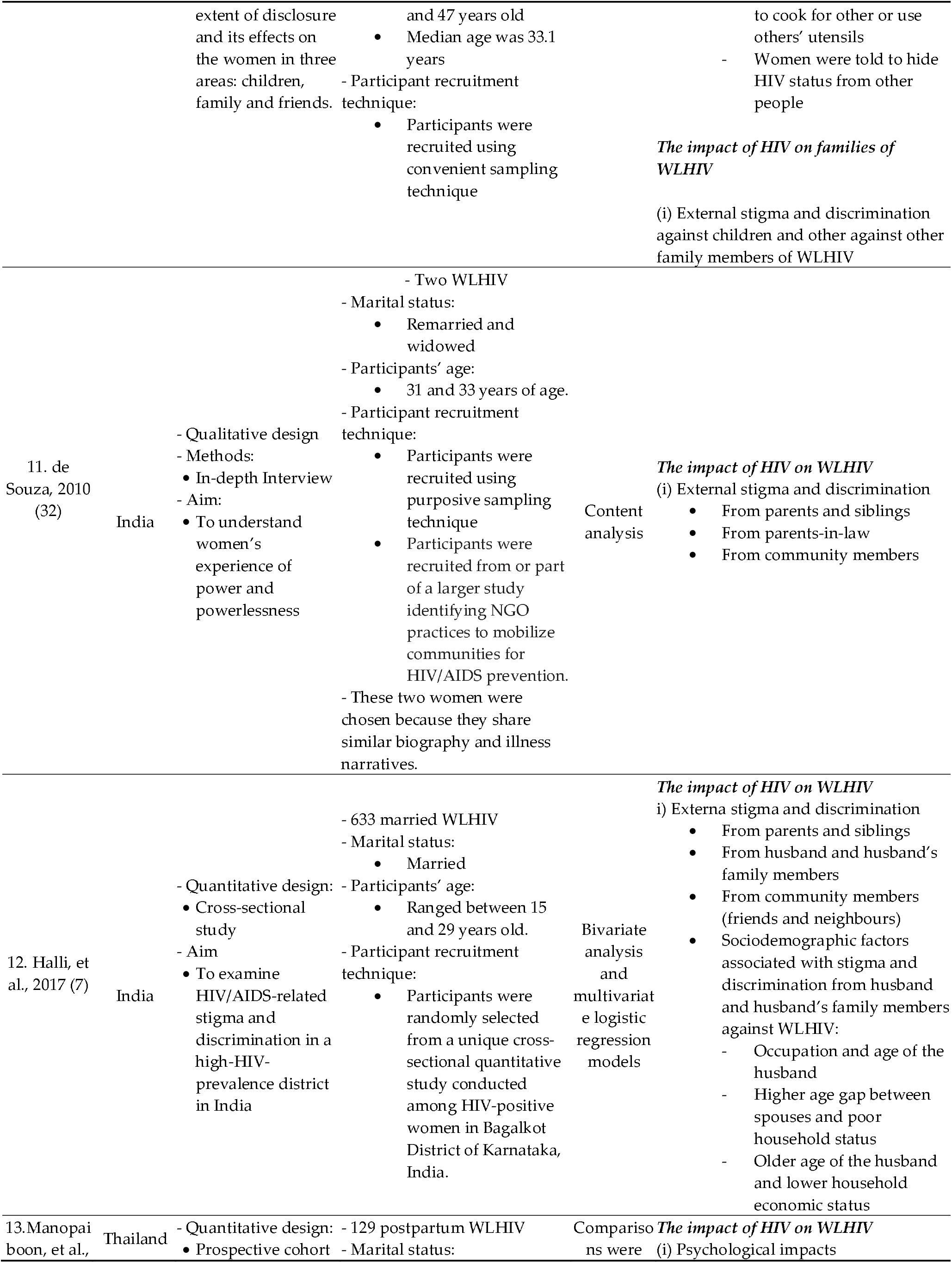
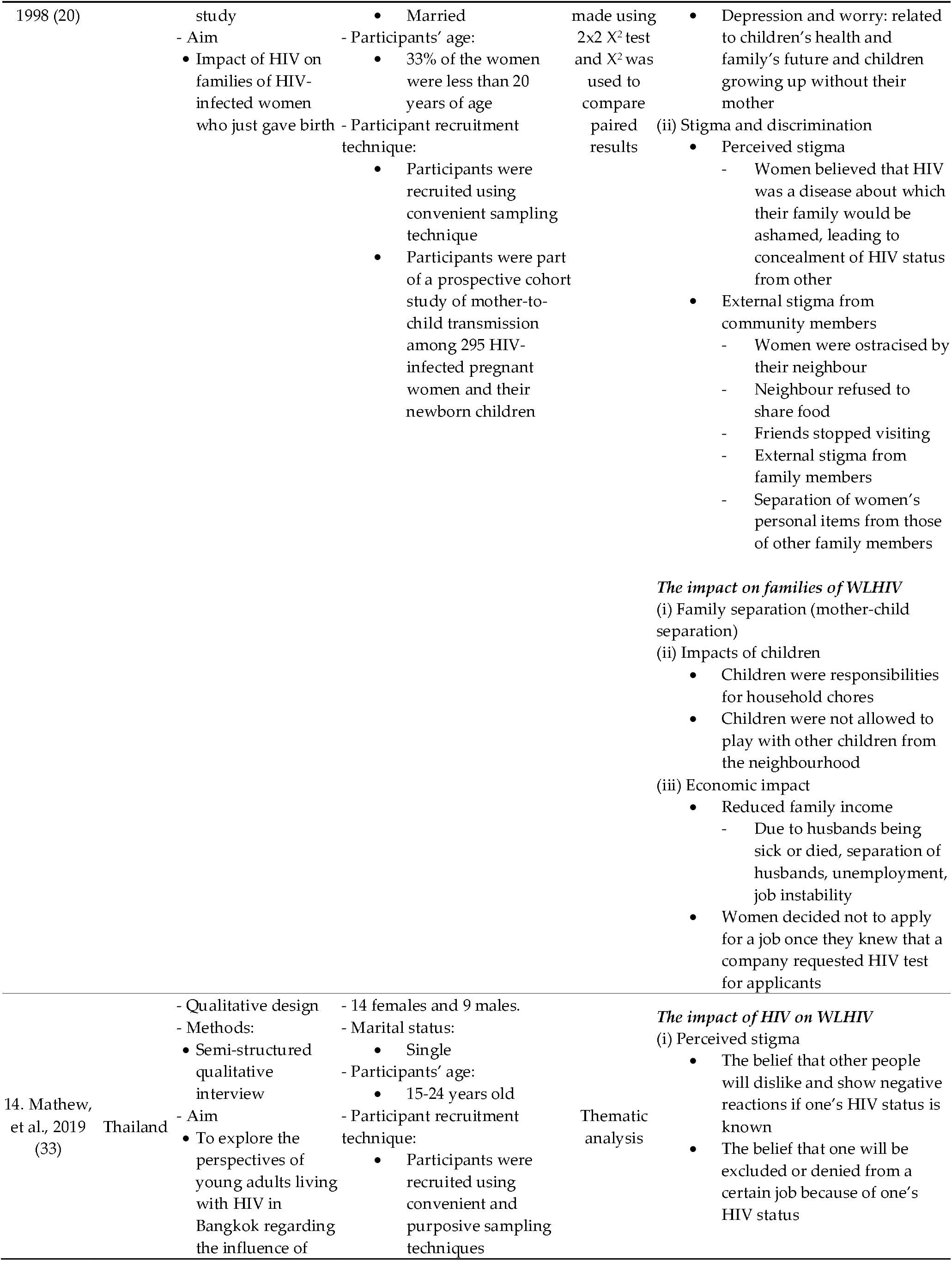
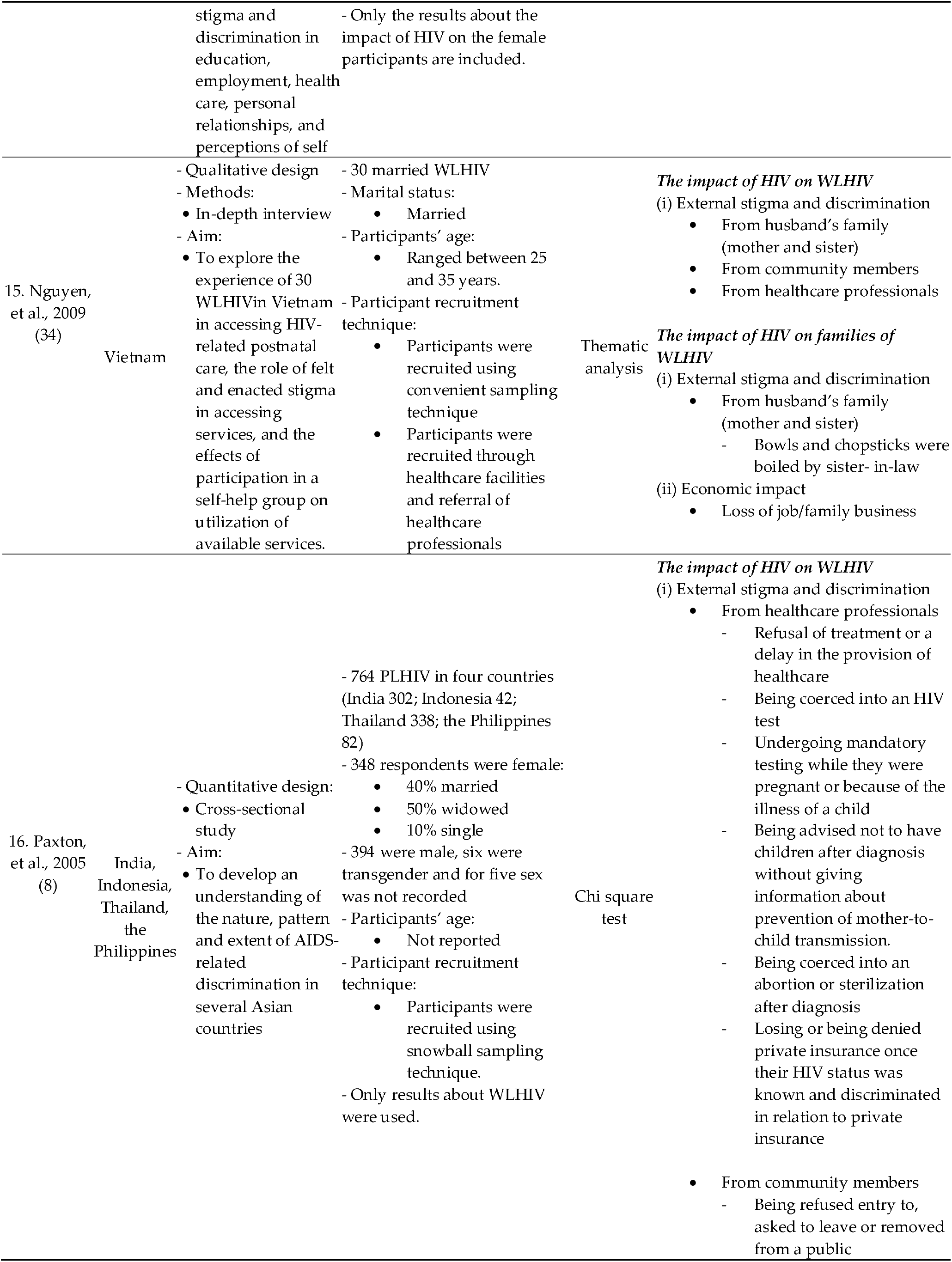
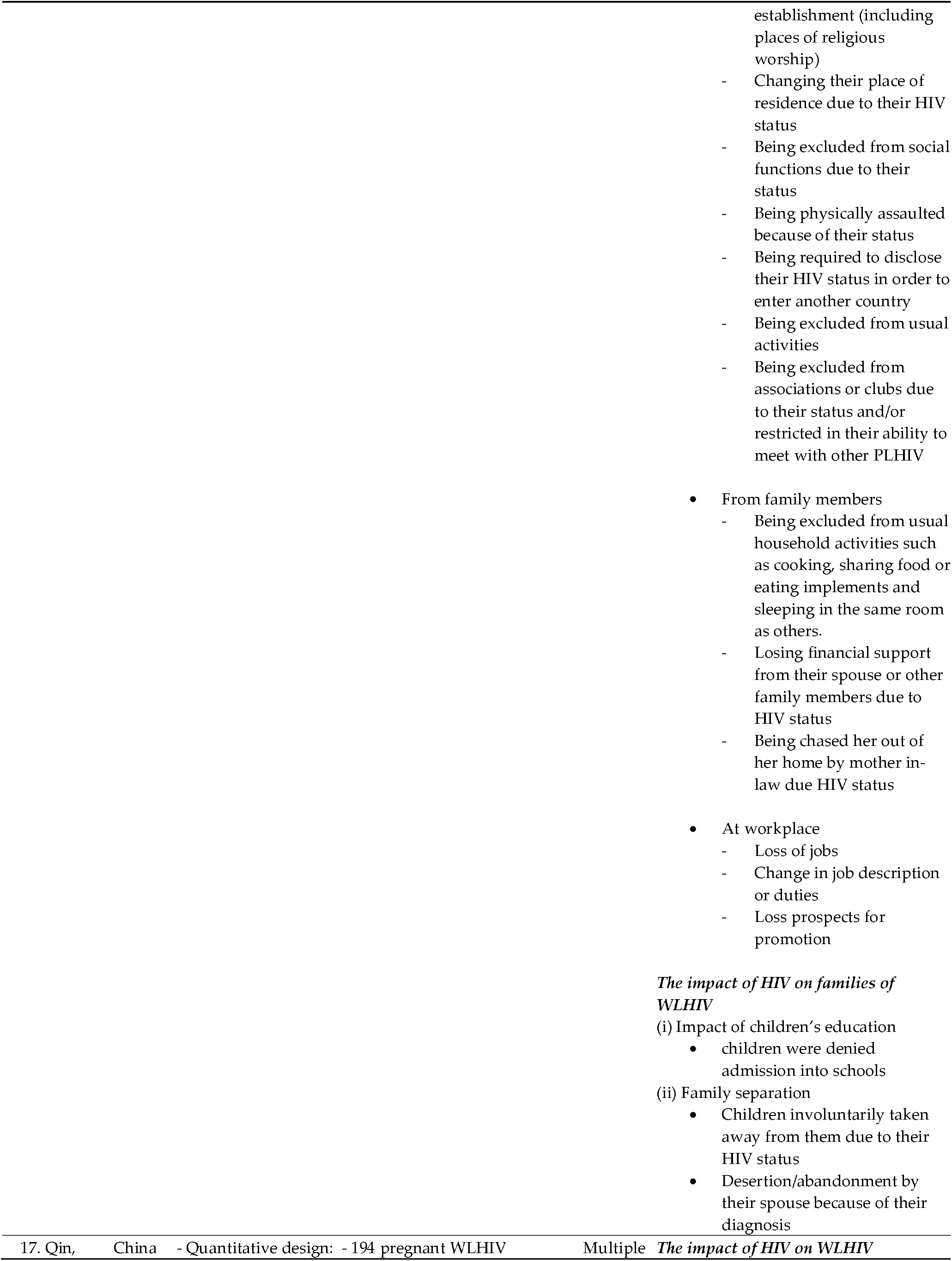
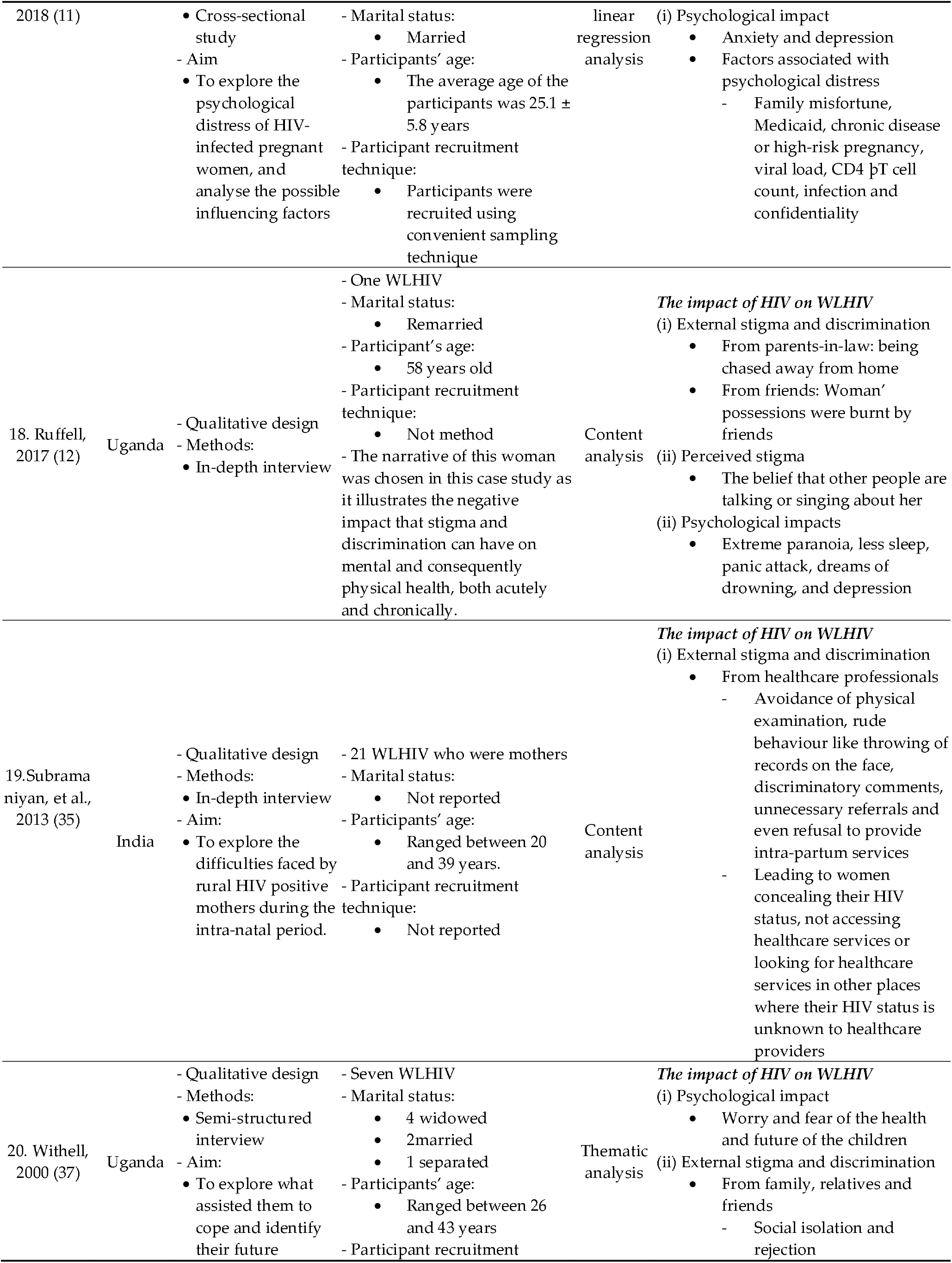
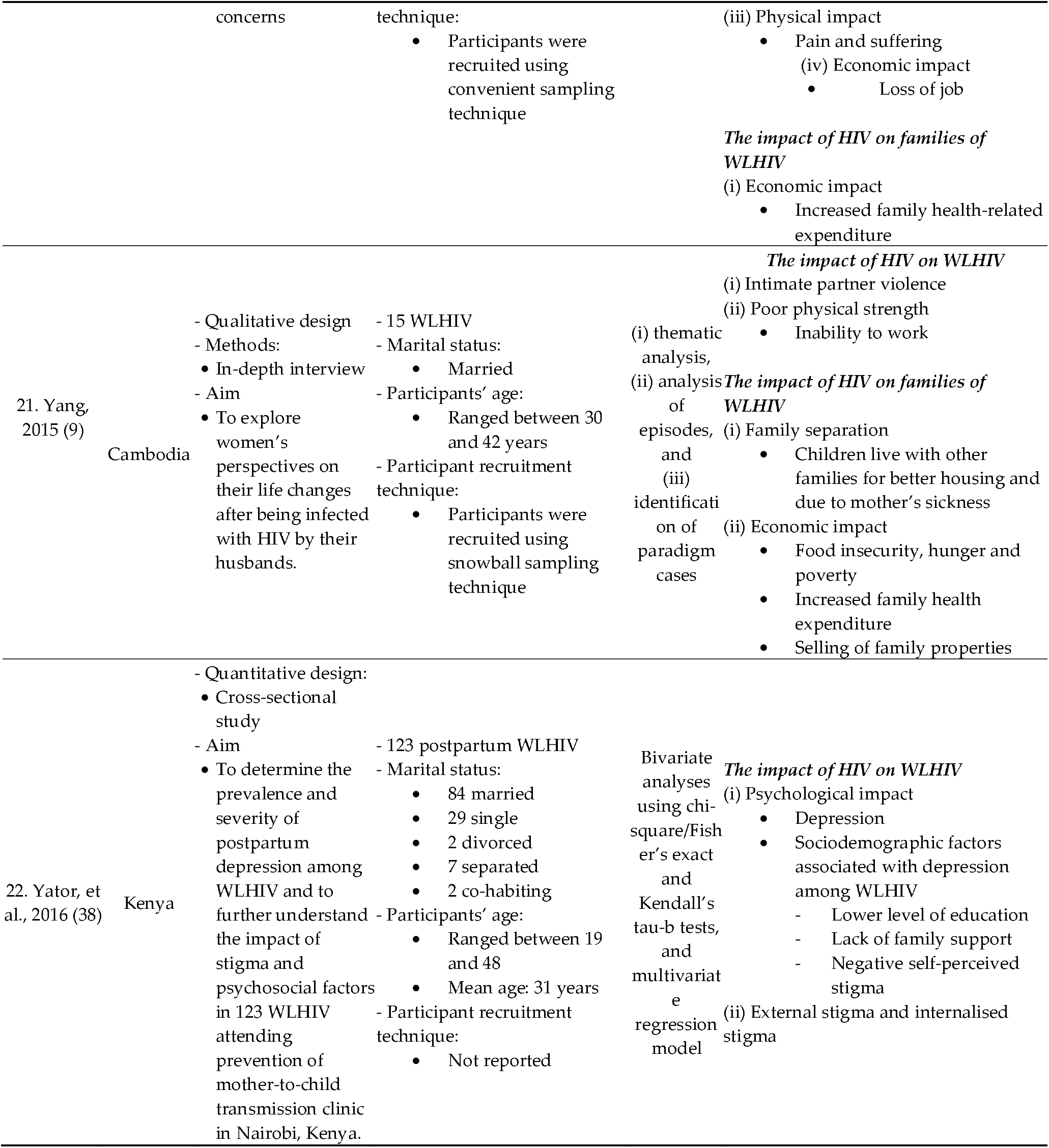
Description of the included studies in alphabetical order.

### 3.2. Impact of HIV on WLHIV and their families

Two thematic categories including individual level and family level impact (Figure 2) were identified. Direct quotes from participants were also selected from the studies to support the descriptive synthesis of each theme as summarised in Table 2. From each of the two categories above, a further synthesis (explanatory phase) was undertaken where different HIV-related impacts were mapped into a comprehensive Conceptual Model of complex impact (Figure 2) endured by WLHIV and their families. These were subcategorised into five thematic areas (Figure 2) including: (i) psychological impact on WLHIV and their families, (ii) physical health impact on WLHIV and intimate partner violence against them, (iii) negative social impact on WLHIV and their families, and (iv) economic impact on WLHIV and their families. These themes are discussed in turn below. The mechanisms through which these influence the lives of WLHIV and their families, and interrelationships between them are also described.

**Table 2.**
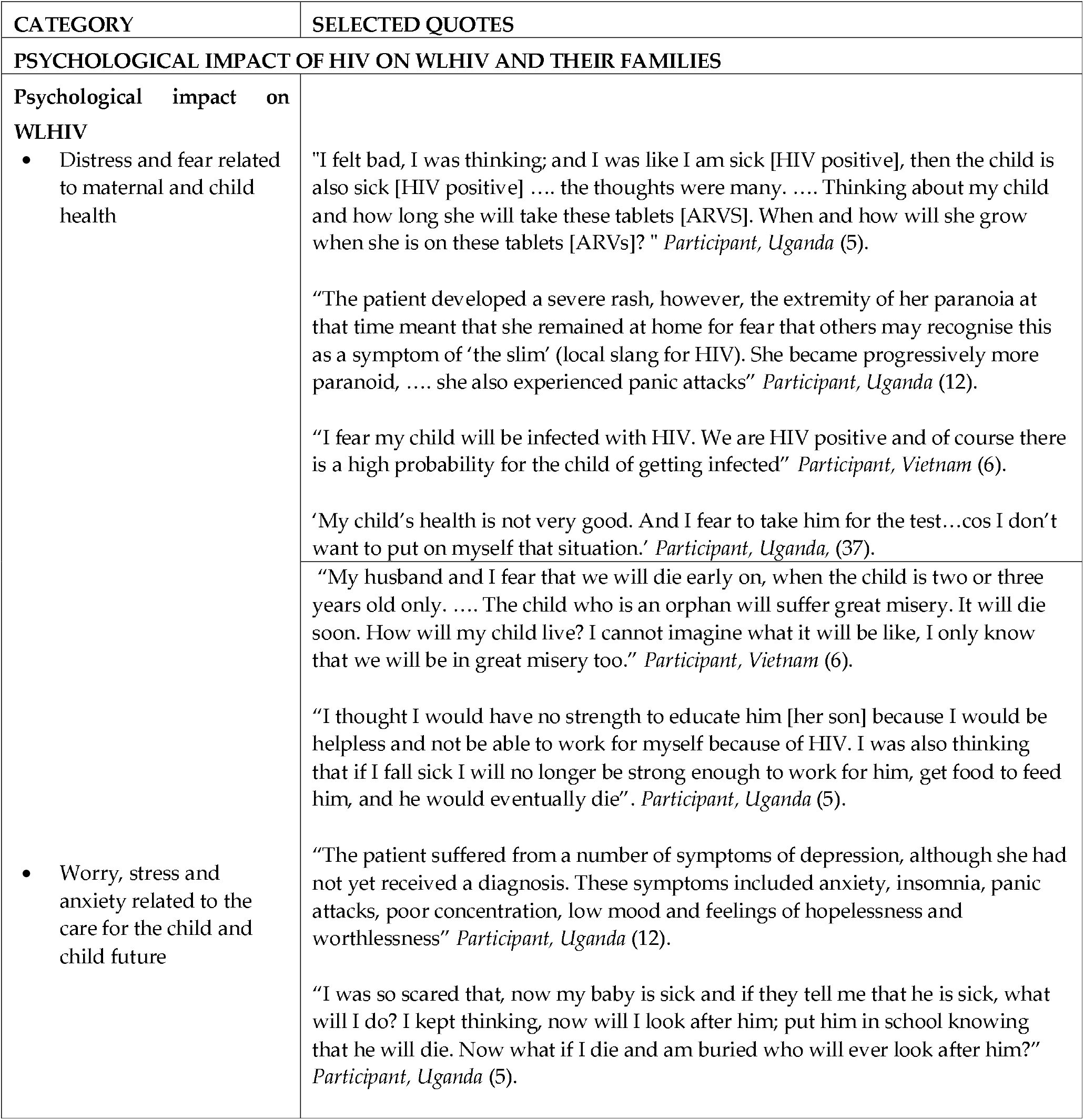
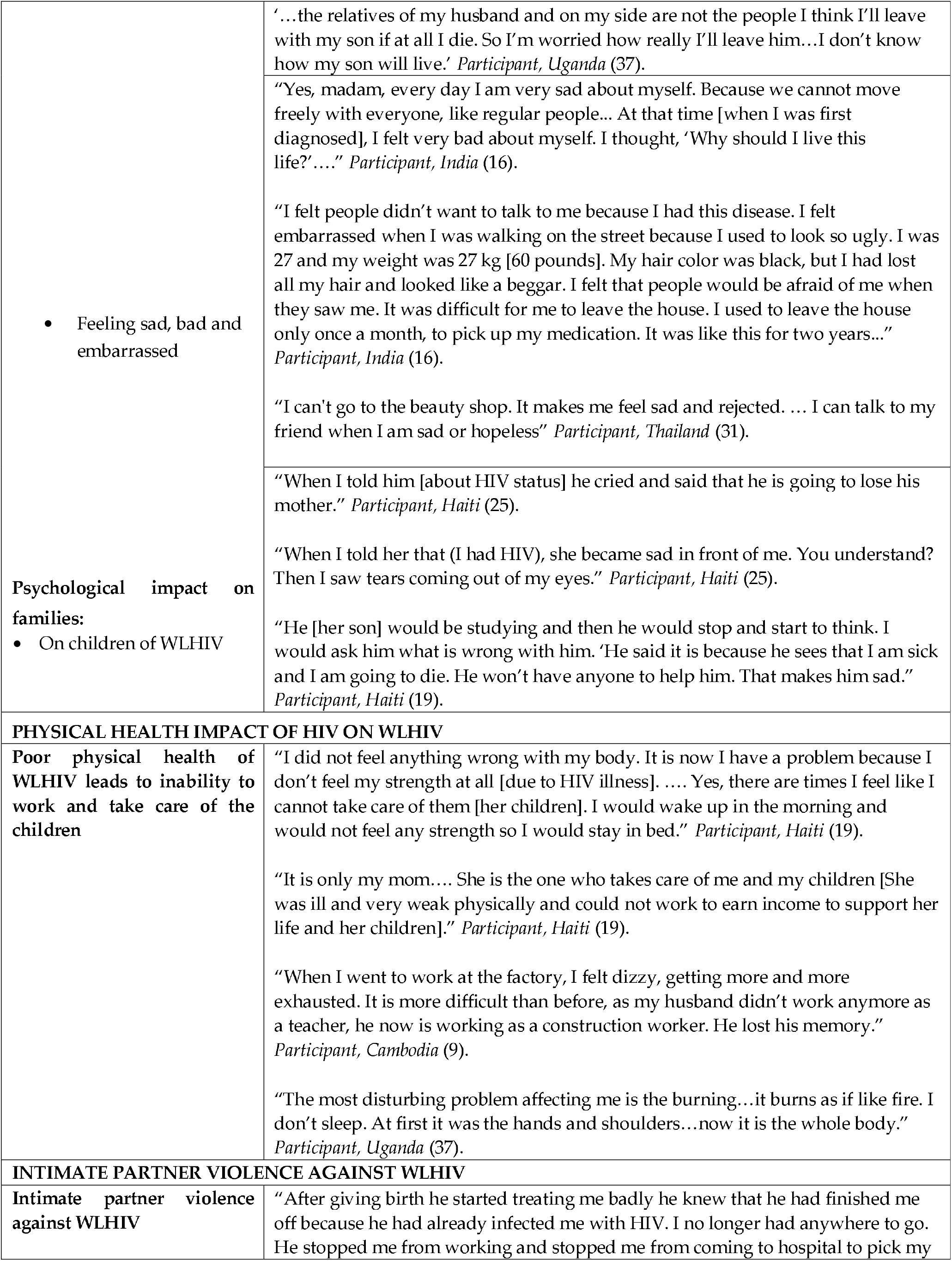
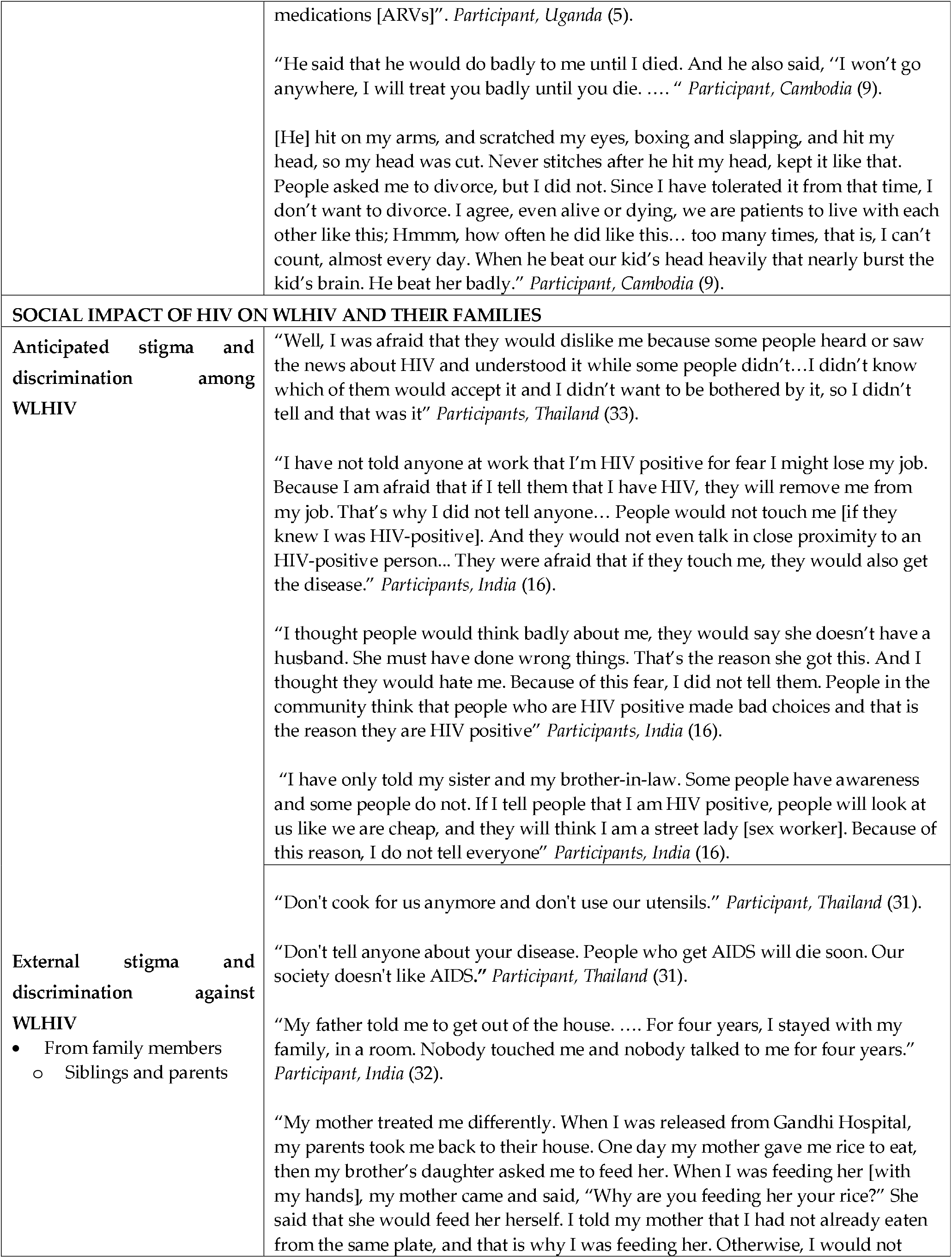
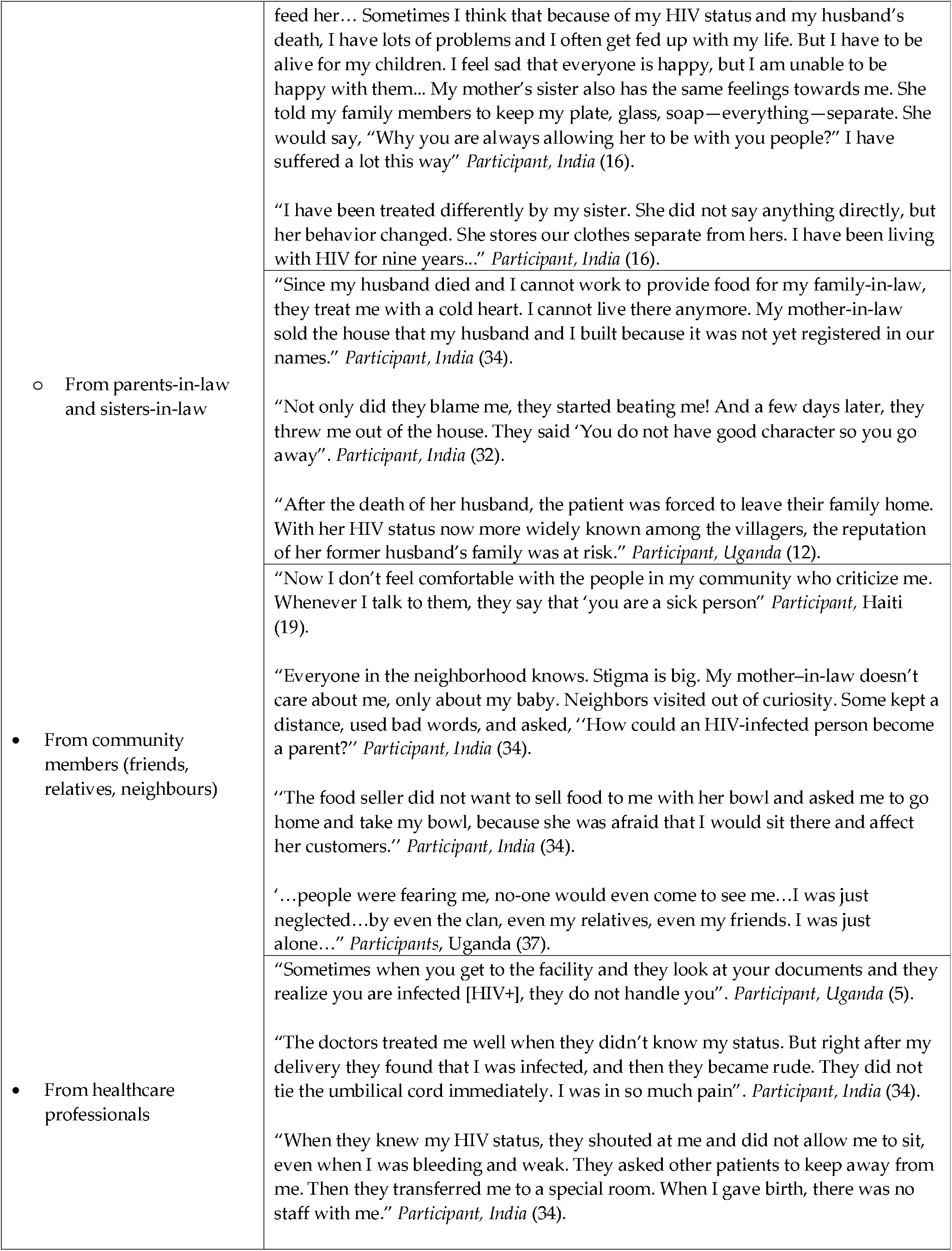
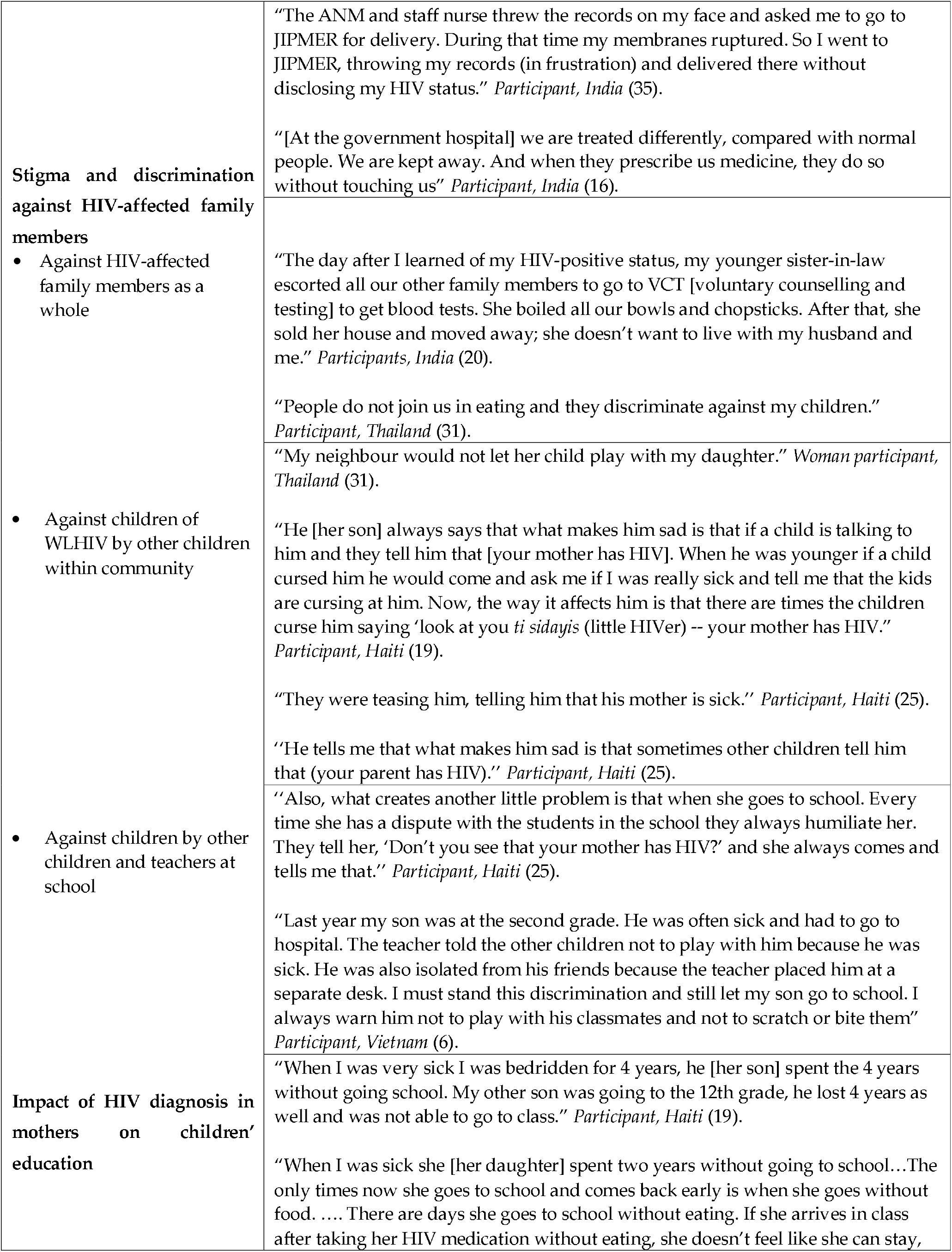
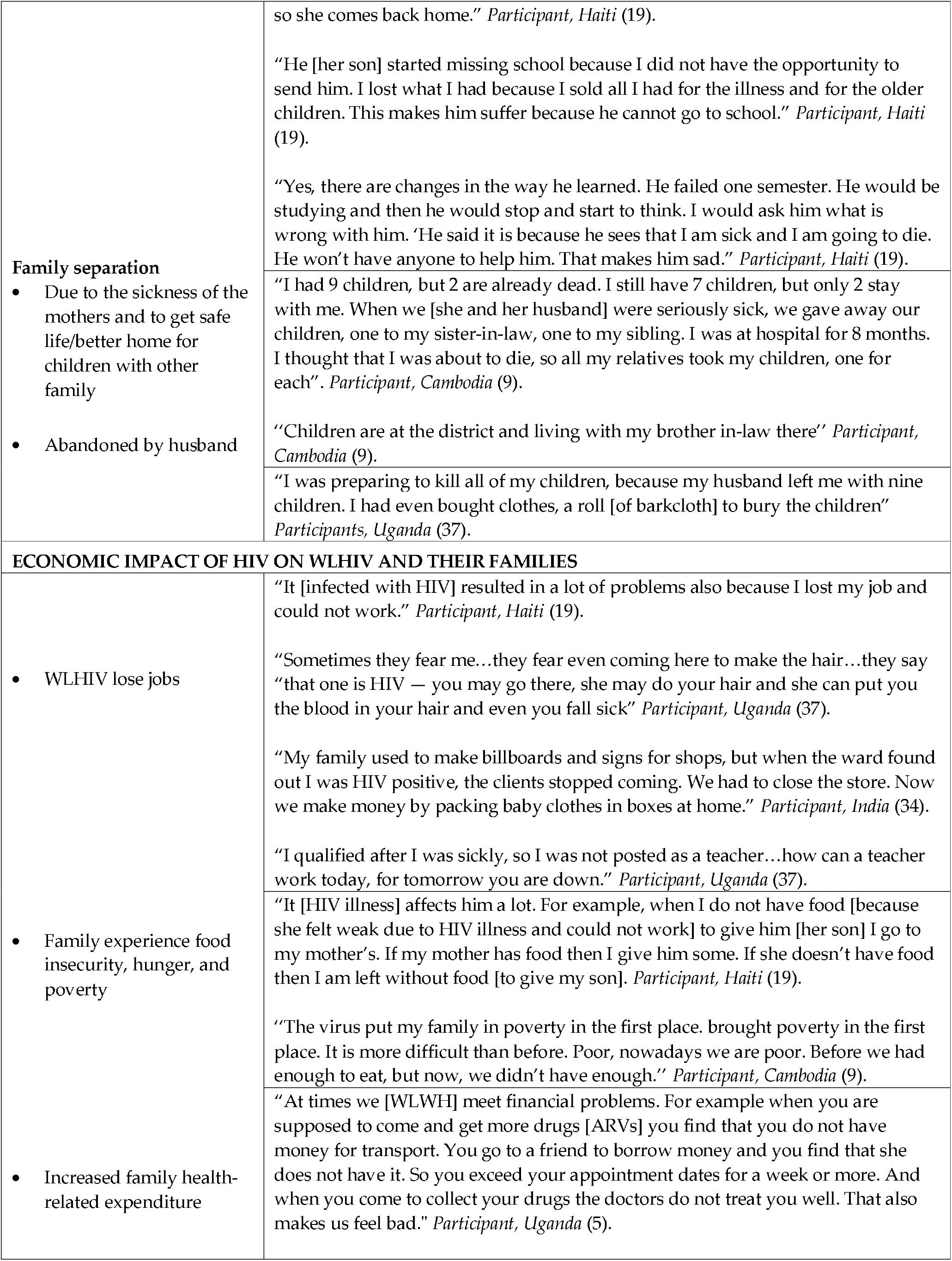
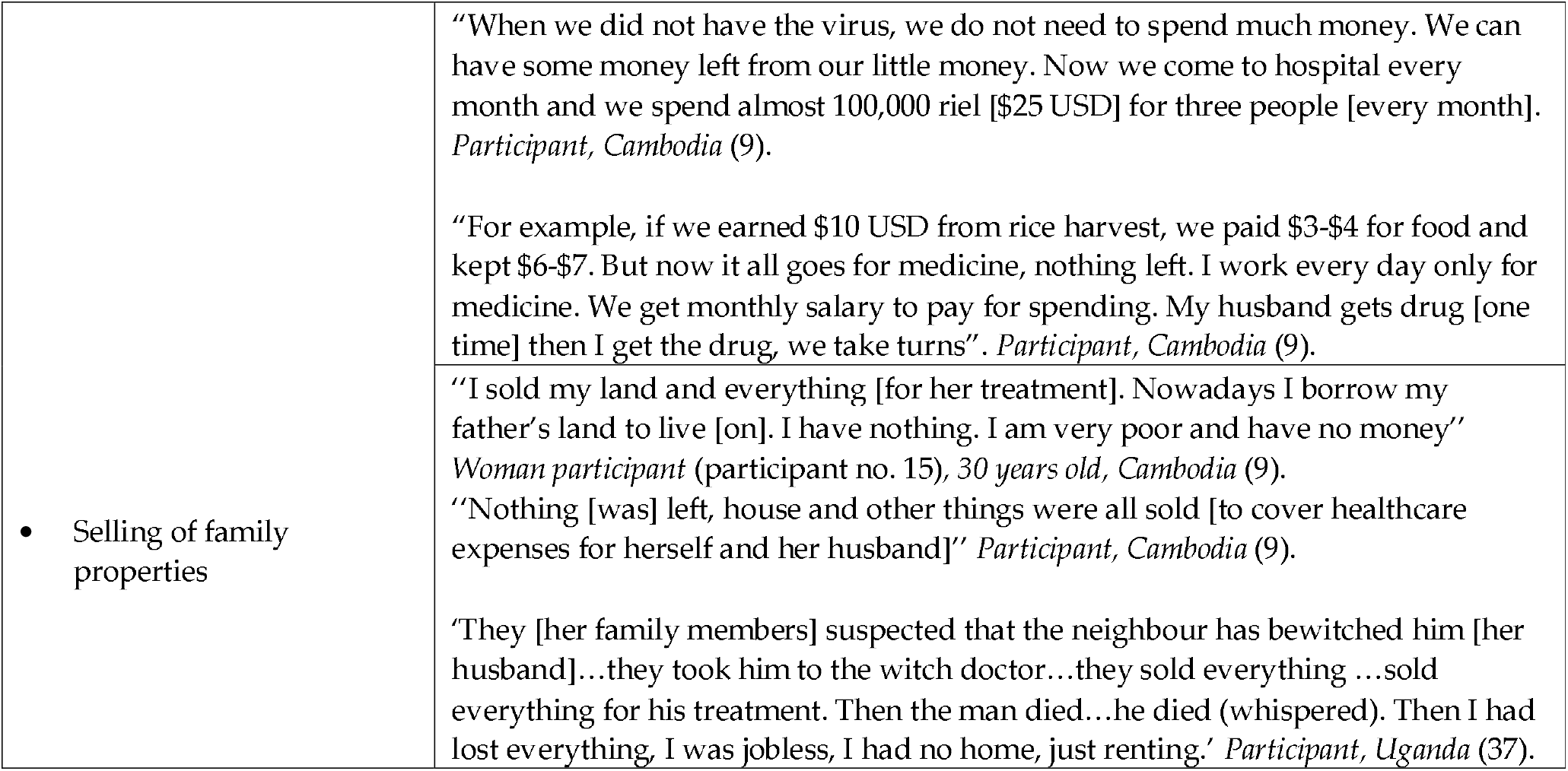
Qualitative evidence of the impact of HIV on WLHIV and their families.

**Figure 2.**
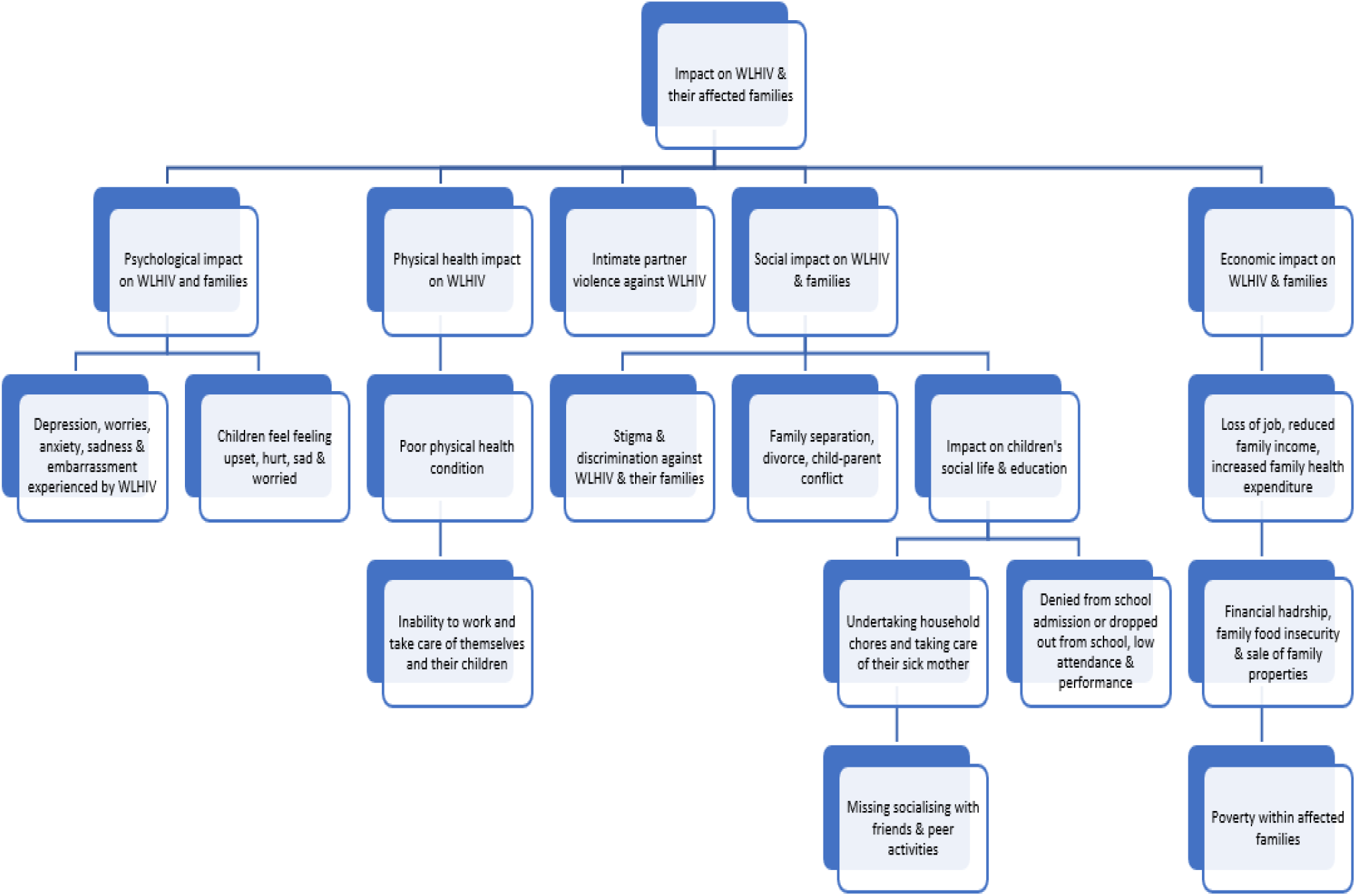
Conceptual Model for HIV impacts on individuals and families.

#### 3.3.1. Psychological impact of HIV on WLHIV and their families

Thirteen studies reported various psychological challenges faced by WLHIV and their families, especially children (5, 6, 11, 12, 16, 19, 20, 25, 30, 31, 36–38). Psychological challenges such as depression, worries, anxiety, sadness and embarrassment were the most common experiences among WLHIV (12, 16). These psychological challenges were often triggered by a range of concerns or factors the women felt or faced following an HIV diagnosis. Moreover, children of WLHIV were psychologically impacted manifesting in a range of emotional responses including sadness and worries about their mother HIV status and poor health condition (19, 25). Advanced stage of HIV infection, fear of breach of confidentiality about HIV status and feeling like a shame to family were also noted as factors associated with the psychological impact on WLHIV (11, 16, 30). Fear of transmitting HIV infection to unborn babies (6), concerns about the future of children upon WLHIV untimely death (5, 6, 20, 37), and the lack of resources to take care of their children (6, 30) put additional psychological stress to WLHIV. The lack of social support, social rejection or isolation, perceived stigma (16, 31, 36, 38) and poor economic conditions due to reduction in work and increase in family healthcare expenses (6, 30) caused depression and increased fear among WLHIV. Additional factors such as lower education attainment, lack of family support (30, 38), being primary caretaker for children, cessation of the relationships with a partner and/or partner’s death (30), poor health, single marital status, unintended pregnancies (36), predicted poor psychological functioning of WLHIV.

#### 3.2.2. Physical health impact of HIV on WLHIV and intimate partner violence (IPV) against them

Three studies reported reduced physical strength (9, 19), difficult sleeping, dizziness and exhaustion (37) after HIV diagnosis among WLHIV. These physical issues were reported to jeopardise the day to day functioning of WLHIV, which affected the way they looked after their children and the ability to work and provide for their children (9, 19).

Two studies reported that a diagnosis of HIV in women results in IPV and maltreatment towards them (5, 9). Husbands’ rage towards wives’ HIV diagnosis, women’s tolerance of husbands’ behaviours and women’s commitment to living together for the sake of children were reported to heighten IPV against WLHIV (5, 9).

#### 3.2.3. Social impact of HIV on WLHIV and their families

##### 3.2.3.1. Stigma and discrimination against WLHIV

Nine studies reported women’s feelings of anticipated stigma following their HIV diagnosis, and they believed that people, including family members, would react negatively to their HIV status (16, 33, 40). For example, they hold the belief that other people will disclose their status and family members will reject them after finding out about the HIV diagnosis (12, 16, 20, 31, 33). Such belief was based on HIV-related discriminatory and stigmatising attitudes and behaviours previously experienced by other PLHIV, and have often led to women isolating themselves to hide the HIV diagnosis from family members (12, 20, 31). A lower perception of social support from other people within their communities (36) and the fear of children being stigmatised (6) were also expressed as a form of anticipated stigma, leading to WLHIV avoiding participation in communal activities. Being older women, having limited education and having a husband with multiple wives (13) were additional factors associated with anticipated stigma.

Nine studies showed that WLHIV experienced HIV-stigma and discrimination from close family members such as husbands and in-laws, parents and siblings. WLHIV experienced blaming, verbal insults, avoidance and rejection from husband and in-laws (7, 16, 32). WLHIV also were accused by parents-in-law of transmitting HIV to their husbands and were expelled from their marital homes (8, 12, 32, 34). Husbands’ occupation, a wider age gap between WLHIV and their husbands, and the women’s lower household economic status, financial dependency on their husband and inability to engage in income generating activities were contributing factors associated with such discriminatory and stigmatising attitudes and behaviours (7). Women’s inability to engage in income generating activities and the financial dependency on their husbands appeared to be an explanation for such negative attitudes and behaviours of husbands and in-laws towards them (20, 34). WLHIV also experienced stigma and discrimination from their own families including from parents and siblings. They were asked to leave home (32, 37), excluded from usual family activities including cooking, and their personal items (e.g., clothes and eating utensils) were separated from those of other family members (7, 8, 16, 20, 31).

Twelve studies reported HIV-stigma and discrimination against WLHIV by friends, neighbours and other community members (5, 7, 8, 12, 13, 19, 20, 31, 32, 34, 35, 37). Social isolation (e.g., refused entry/excluded from social functions or removed from public establishments) (8, 13, 37), refusal of neighbours and relatives to share food and drinks (20, 31), having personal possessions burned by friends and relatives, and eviction from rental properties (12) due to the fear of contracting HIV through social contacts, were some discrimination incidents that WLHIV faced within communities. WLHIV also experienced physical assaults, negative labelling using discriminatory words such as “HIV carriers” or “she is (HIV) positive”, and harassments by other community members (8, 32, 34).

Similarly, within health care settings, WLHIV experienced a range of discriminatory treatments or behaviours by healthcare professionals due to limited knowledge about HIV and fear of contracting HIV (5, 8, 13, 16, 34, 35). Health professionals exerted the HIV-stigma and discrimination towards WLHIV in a form of criticising, blaming, shouting at, throwing health records in their face (5, 34, 35), avoiding physical contact with them (34, 35), refusing to provide healthcare services or leaving them untreated (5, 8, 13, 16, 34, 35), and unnecessary referrals (16, 35). Coercion to undergo HIV testing, termination of pregnancy, sterilisation, and termination or loss of private health insurance, after HIV diagnosis, were other instances of healthcare-related discrimination against WLHIV (8). As a consequence, WLHIV chose not to disclose their HIV status and would go untreated (34, 35), often leading to advanced stage of HIV infection.

Stigma and discrimination towards WLHIV also occurred within the workplace settings, manifesting in the loss of prospects for promotion and unexplained changes in job descriptions due to employers worrying that customers may avoid using the services provided by WLHIV (8, 37).

##### 3.2.3.2. Stigma and discrimination against HIV-affected family members

Five studies reported that husbands and children of WLHIV, who were HIV-negative, also experienced stigma and discrimination from relatives and other community members (6, 19, 25, 31, 34). For example, husbands’ relatives sterilised (by boiling) utensils (e.g., bowls and chopsticks) that had been used by the women’s husband and children before they could use these themselves due to the fear of contracting HIV, stemming from a lack of HIV knowledge (34). Children of WLHIV were also negatively affected by their mother’s HIV-positive status.

##### 3.2.3.3. Impacts of mothers’ HIV status on children’s social life and education

Seven studies indicated that children of WLHIV experienced social and educational challenges (8, 19, 20, 25, 31, 39, 41). Children of WLHIV often faced stigma and discrimination, such as being rejected, teased or mocked by their friends and significant others due to their mothers being diagnosed with HIV infection. For example, they were told “your mother has HIV”, called “little HIVer” by friends (19, 25) and placed at a separate desk by teachers, while other children were told not to play with them at school and within community where they lived (31, 41). Two studies reported that because of their mother’s regular illness, children of WLHIV had additional household responsibilities including undertaking household chores and taking care of their sick mothers (20, 39). These additional responsibilities prevented children from activities such as socialising with friends, being involved in extracurricular school or peer activities and doing school homework (20). Mothers’ poor health condition due to HIV infection also influenced children’s education through missing school and poor availability of essential provisions such as food and school fees, leading to children’s poor educational performance and/or outcomes (19). In one study, children of WLHIV were denied school admission or expelled from school due to pressure or complaints from the parents of other students, who feared HIV transmission to their children (8).

##### 3.2.3.4. Family separation and child-parent conflicts

Forced removal of children by in-laws due to a mother’s HIV diagnosis was another social impact of HIV on HIV-affected families (8, 20, 34). WLHIV also reported voluntarily sending their children to live with their siblings or sisters-in-law/ brothers-in-law’s family, once they were terminally ill or hospitalised (9). The main reason for such separations was due to the fear of in-laws about mother-to-child transmission and the mothers’ inability to raise their children due to poor physical health and economic conditions (9, 19). Husband-wife separation or abandonment by a husband or partner was another negative impact of HIV on the families of WLHIV (8, 19, 37), making it hard for women to take care of themselves and their children (37). A diagnosis of HIV in mothers also brought conflicts to parent-child relationships (39), and in some instances, children blamed their fathers as the source of their mothers’ HIV infection (25). This all would likely to be worse if the children were HIV-infected themselves (39).

#### 3.2.4. Economic impact of HIV on WLHIV and their families

Six studies reported that HIV infection had negative economic impact on WLHIV at individual and family levels, through several mechanisms. For example, a loss of job or potential work due to women’s HIV status (8, 37) and poor physical strength (9, 37) were reported to prevent them from working, affecting their economic situation. HIV diagnosis negatively influenced women’s willingness to apply for jobs that required HIV testing (20) and led to women losing financial support from their spouse and other family members (8).

HIV impact at a family level included the reduction in family income due loss of job or unemployment and women’s inability to work (20), and in some cases clients or customers stopped using the family business services after learning about the HIV status of women in the family (34), severely impacting the family business. An HIV diagnosis in women can also increase family healthcare expenses (9, 37), necessitate forced sale of family properties (including land and houses) to cover healthcare and living expenses (9, 19), putting additional economic strain on families. This leaves the family in a vicious cycle of disadvantages including food insecurity and poverty (9, 19). Table 2 below summarises the qualitative evidence of the impacts of HIV on WLHIV and their families.

## 4. Discussion

### 4.1. Summary of findings

The primary objective of this review was to understand the impact of HIV on WLHIV and their families in LMICs. A total of 22 studies comprising of nine quantitative and 13 qualitative studies were included in this review. Six quantitative (7, 11, 13, 30, 36, 38) and four qualitative studies (5, 12, 32, 35) focused on exploring the impact of HIV on WLHIV, while two quantitative (8, 20) and six qualitative (6, 9, 19, 31, 34, 37) studies looked at the impact on WLHIV and their families. One quantitative (39) and qualitative (25) study each only investigated the impact on families of WLHIV.

Psychological challenges faced by WLHIV were presented in five quantitative studies (11, 20, 30, 36, 38), six qualitative studies (5, 6, 12, 16, 31, 37), while two qualitative studies also reported psychological impact on the women’s children (25) and on both the women and their children (19). Poor physical health impact on WLHIV (9, 19, 37) and IPV against them following HIV diagnosis (5, 9) were reported in these qualitative studies. Negative social consequences of an HIV diagnosis in women, such as stigma and discrimination against them within family, community, workplace and healthcare settings were reported in six quantitative studies (7, 8, 13, 20, 36, 38), and 10 qualitative studies (5, 6, 9, 12, 19, 31, 32, 34, 35, 37), while stigma and discrimination towards their family members, especially children within family, community, healthcare and school settings were reported in five qualitative studies (6, 19, 25, 31, 34). Other negative social consequences on the women’s families, such as family separation, child-parent conflict, HIV impact on children’s education and social life were reported in two qualitative (9, 19) and three quantitative (8, 20, 39) studies. Negative economic impact experienced by WLHIV (37) and their families (9, 19, 34, 37) were presented in five qualitative studies. One quantitative study also reported economic impact on family of WLHIV (20).

### 4.2. An explanatory conceptual model for the impact of HIV on WLHIV and their families

Two levels of HIV impact were identified: the impact on WLHIV at an individual level and on families affected by the women’s HIV diagnosis. Drawing on the reviewed studies and the conceptual framework for the socio-economic impact of the HIV/AIDS epidemic on households (42), an explanatory Conceptual Model for HIV impact on WLHIV and their families was developed to visually represent the connection between the emerging themes (see Figure. 2).

The Model demonstrates that HIV infection results in a range of negative consequences at the individual and familial level. At the individual level, HIV infection causes psychological challenges including depression, apprehension and anxiety on WLHIV (11, 12, 43), often triggered by concerns that the women may face after being diagnosed with HIV. These may include concerns about their poor health condition, HIV status, stigma, discrimination, family shaming and the future of their children (11, 16, 30). Such concerns reflect psychological responses of WLHIV towards the knowledge about their HIV-positive status and the possibility of various negative consequences that may occur to themselves or their families (44). Similarly, HIV infection can cause psychological challenges on the women’ families, especially children, such as feeling upset, hurt, sad and worried about the mother poor health condition or HIV status (19, 25). HIV can also lead to poor physical strength which influences the ability of WLHIV to work and take care of themselves and their children effectively (9, 19, 37). This can lead to the women being dependent on the support from their extended families to fulfil their basic necessities and to take care of their children, which are often burdensome for families, especially the ones with limited resources (9, 19). Abuse of WLHIV in any forms (e.g., physical, verbal, sexual abuse) by significant others including husbands, partners and in-laws due to unacceptance of women’s HIV status has been reported in many LMICs, further exacerbated by sociocultural and patriarchal systems and structures that require wives’ submission to husbands or male partners (45–49). For example, in some cultures in Uganda, Zimbabwe, India and Cambodia, husbands are deemed to submit to husbands (46–50). Non-submission to husbands would result in sanctions, including beating, divorced, chased from home and not financially supported (46).

HIV infection can also lead to negative social impact, such as stigma and discrimination against WLHIV, reflected in a range of negative attitudes and behaviours towards them, including rejection, insults, avoidance and isolation, which may be imposed by family members, community members, healthcare professionals, and employers or colleagues (7, 12, 13). A diagnosis of HIV in women can also lead to courtesy stigma towards their family members, especially children, manifesting in rejection or negative treatments by their friends and significant others within communities or school settings (8, 19, 25, 31, 41, 51). Stigma and discrimination towards the women themselves and also their family, are often very damaging and in many cases lead to feelings of rejection, hopelessness, shame, anger, and fear (11, 12, 16, 20, 31). Notably, the lack of knowledge of HIV transmission and prevention that is reflected in the fear of acquiring HIV infection through interaction with PLHIV is one of the biggest sources of HIV stigma and discrimination (8, 15, 52). Stigma and discrimination towards WLHIV and their family seem to also reflect psychological response of non-infected people towards the knowledge about the existence of PLHIV around them, who may transmit the infection to them or threaten their health and life (17, 18, 44).

Denial of school admission and/or withdrawal of children from school due to mothers’ inability to pay school fees and provide children’s basic necessities, discrimination, and children’s involvement in home duties due to poor health of WLHIV, are also serious social consequences of HIV on children of WLHIV (42, 53–55). Children of WLHIV are an at-risk population, they often miss out on crucial socialisation with friends and extracurricular activities due to increased responsibility at home (8, 19, 20). These are educational consequences that hinder opportunities for their future, but are often in conjunction with feeling upset, hurt and worried about their mother’s poor health, and/or being subjected to discrimination (8, 19). These children are also at high risk of mental health issues, often without any sort of support in place (56, 57). Forced and voluntary family separations of child-parent or husband-wife, have also been reported, either due to poor physical health, inability of WLHIV to take care of their children, lack of support, or unacceptance of women’s HIV status by husband or partner (8, 9, 19, 37). The lack of support and unacceptance of women HIV status, especially by people around them, such as families and friends, can lead to self-isolation, lack of access to health care and treatment services and inability to cope with various challenges facing them (11, 31, 58, 59).

Consistent with the constructs of the framework for the socio-economic impact of HIV/AIDS on households (42, 53, 60–62), reduced family income due to loss of employment of infected family members (WLHIV or their partners/other family members), or loss of a family business, in addition to increased family health expenditure, leads to economic or financial difficulties (8, 9, 37). These economic consequences may also lead to household food insecurity prompting decisions such as selling of family properties (e.g., land and houses), leading to a vicious cycle of poverty and cascade of disadvantage within HIV-affected families (60–63). This chain of economic consequences can further lead to deterioration of WLHIV’s physical and psychological health (21–23).

### 4.3. Implications for future practices or interventions

This systematic review summarises evidence on a range of HIV negative consequences on an entire family unit when a woman is diagnosed with HIV in a LMIC. It highlights psychological, physical, social and economic aspects that need to be addressed in future responses.

#### 4.3.1. Addressing stigma and discrimination against WLHIV and their children

HIV is a stigmatising infection, which varies according to culture and context (64). Many cultures associate HIV with immorality through an association of the disease with groups deemed ‘deviant’ from social norms, such as homosexual men, commercial sex workers and drug users (65). In this review, though not obvious that WLHIV were linked with any deviant behaviours, their experience of stigma and discrimination was within family, community, healthcare and workplace settings. Children of WLHIV were also stigmatised and discriminated against by their friends and teachers at school, and by other community members, just for being related to someone with HIV. These appeared to be influenced by people’s assumption and fear that children of WLHIV could also be HIV-infected themselves and could transmit the infection to other children (17, 18, 44) To respond to HIV-stigma and discrimination effectively, interventions should target not only at individual or family levels, but also at community and societal levels, including family, community, school, healthcare and workplace settings.

HIV education and awareness of its transmission methods for population groups, families and community members would be a vital component of any intervention, seeking to raise awareness of HIV/AIDS as well as acceptance of WLHIV (66). The aim of this would be to reduce stigma and discrimination against WLHIV and allow their family and community to provide social support (66–68). The key stakeholders in the community would be critical in supporting any HIV intervention, allowing them to lead the community through the stages of the intervention (69). Their opinions and values matter (69). Similarly, educating and involving both PLHIV who have had their viral load suppressed and been open about their HIV status, and healthcare professionals who provide care to PLHIV, in the delivery of such an intervention would be equally important. They could reinforce the messaging regarding HIV transmission and safety measures to be adopted. Involvement of these people may bring stronger influence on community members and increase the likelihood of community members receiving the intervention, which can lead to better acceptance of WLHIV and other PLHIV in general and reduction or elimination of stigma and discrimination towards them within families, communities and other settings (70–73). Health systems interventions, such as intervention supporting HIV patient self-management and improving patient engagement in HIV care can also be useful strategies to build positive self-perceptions and confidentiality of WLHIV and to address stigma and discrimination against WLHIV (74, 75). These may also lead to reducing internalised stigma which often comes from negative self-perceptions, over worried and fear of other people’s reactions.

#### 4.3.2. Addressing psychological challenges on WLHIV

Despite improvements in knowledge about HIV and increased access to HIV treatment and support across the world (1), an HIV diagnosis is still found to impose significant psychological challenges on WLHIV, manifested in a range of emotional responses, including sadness, apprehension and anxiety (1, 5, 16, 38). As such, interventions are needed that tackle psychological challenges faced by WLHIV and their families (76). Counselling, provided by a trained HIV-counsellor, could be a useful strategy where WLHIV could receive psychosocial support once they are diagnosed with HIV (77). Counselling, in its very nature involves sharing personal experiences and feelings, and therefore counselling services need to consider and account for sex, religious and cultural aspects or norms and values of WLHIV which may increase acceptance and access to the services (78, 79). Family counselling could be a great benefit for WLHIV as it can increase their understanding and acceptance of the infection, reduce stigma and improve treatment of WLHIV (68, 80, 81). Counselling is also reported to increase patients’ adherence to ART which can improve their physical health condition (82). An additional strategy to support WLHIV would be to provide HIV peer support groups, providing them with opportunities to share information, experiences, and gain support from others (70, 72, 73).

#### 4.3.3. Addressing economic impact of HIV on WLHIV and their families

To address the economic impact of on HIV on WLHIV and their families, some strategies that could be used are the provision of skill training based on WLHIV’ interests and start-up capital for their own business, which have been reported elsewhere effective in supporting economic empowerment and reducing HIV vulnerability among PLHIV (83–86). The provision of training and start-up capital for WLHIV can be done in collaboration between government and other sectors such as NGOs that can provide continuous support, guidance and supervision. In addition, HIV interventions that consider a broader context of women’s lives, including their responsibility for household and caring role within family are also recommended (83). Furthermore, there has to be free access to HIV-related healthcare services for WLHIV which would definitely reduce financial burden or expenses for health care (87, 88). It is also important that much more needs to be done, particularly in improving the quality and access of services to ease the burden of healthcare expenses through provision of free or subsidised health care insurance. An example of such strategy has been implemented in Indonesia, where PLHIV are equipped with health insurance provided by government, which enables them to have free access to healthcare services (89).

### 4.4. Implication for future studies

None of the reviewed studies focused on psychological impact of HIV on HIV-affected family members of WLHIV, though some studies reported scant information related to psychological impact on children. Similarly, no studies reviewed looked at stigma and discrimination against family members other than children, nor explored the impact of women’s HIV status on the employment of family members. The role that cultural and religious values, norms and thoughts play in exacerbating the impact of HIV on WLHIV and their families in LMICs was also a topic not covered. Also, none of the studies reviewed explored the impact of the women’s HIV-positive status on their access to healthcare services or ART. Future studies that address all of the aspects are recommended in order to develop specific and effective interventions tailored to specific environment and settings, for both WLHIV and their family members. For example, findings of future studies that explore the impact of HIV on the women’s access to ART can be used to improve HIV-related healthcare system, tailor HIV care service delivery to address the need of PLHIV and support their health and wellbeing.

### 4.5. Limitation of the study

This review only included studies conducted in LMICs. Thus, evidence on the lived experiences of WLHIV in developed countries about the impact of HIV on themselves and their families was not covered in this review. To build a comprehensive picture of HIV impact facing WLHIV and their families, future literature reviews that include studies on this topic conducted in developed countries are also recommended. Another limitation is that this review included English peer review literature only, thus may have missed studies on this topic, which are reported in other languages.

## 5. Conclusions

The Conceptual Model shown in this review presents evidence that an HIV diagnosis brings a number of significant, persistent, and negative consequences for WLHIV and their families. This Model may assist in designing interventions that address psychological, physical, IPV, social and economic needs to ensure that WLHIV and their families have the best quality of life, and the support they need to thrive even in the face of a HIV diagnosis.

## Supporting information

Supplemental PRISMA checklist

## Data Availability

All data produced in the present work are contained in the manuscript

## Ethics approval and consent to participate

Not applicable

## Data availability

All data generated or analysed during this study are included in this published article

## Acknowledgments

Not applicable

## Supplementary Materials

Appendix 1: Full searching strategy by databases, Appendix 2: JBI Critical Appraisal instruments, Appendix 3: Assessment of methodological quality, PRISMA 2009 checklist: Preferred Reporting Items for Systematic Reviews and Meta-Analyses

