## Supplemental appendices for "The impact of HIV on women living with HIV and their families in low- and middle-income countries: A systematic review"

**Appendix 1:** **Full searching strategy by databases**

Database(s): **Ovid Emcare**
Search Strategy:

| **#** | **Searches** | **Results** |
| --- | --- | --- |
| 1 | human immunodeficiency virus/ | 21298 |
| 2 | acquired immune deficiency syndrome/ | 77403 |
| 3 | (HIV* or "Human immunodeficiency virus" or AIDS).tw,kw. | 438274 |
| 4 | 1 or 2 or 3 | 452954 |
| 5 | female/ or marriage/ or spouse/ or wife/ | 9399966 |
| 6 | mother/ or adolescent mother/ or expectant mother/ or mother child relation/ | 66053 |
| 7 | female/ or female by marital status/ or girl/ | 9394650 |
| 8 | (Wives or Wife or Mothers or female* or girl* or wom?n).tw,kw. | 2455822 |
| 9 | 5 or 6 or 7 or 8 | 9818864 |
| 10 | risk factor/ | 917105 |
| 11 | sexual behavior/ or casual sex/ or concurrent sexual partnership/ or contraceptive behavior/ or extramarital sex/ or hiv serosorting/ or premarital sex/ or prostitution/ or sexual practice/ or sexual promiscuity/ or sexual violence/ or transactional sex/ or unsafe sex/ | 86054 |
| 12 | condom/ | 10909 |
| 13 | social aspect/ | 25625 |
| 14 | peer pressure/ | 768 |
| 15 | social norm/ | 1805 |
| 16 | cultural factor/ | 23440 |
| 17 | socioeconomics/ or economic aspect/ or educational status/ or lowest income group/ or poverty/ | 95665 |
| 18 | social interaction/ | 1124 |
| 19 | social environment/ or environmental factor/ | 44076 |
| 20 | social stigma/ or stigma/ | 11233 |
| 21 | social discrimination/ or social problem/ or employment discrimination/ or social distance/ | 9750 |
| 22 | psychological aspect/ | 53200 |
| 23 | health/ or child health/ or family health/ or mental health/ or sexual health/ or women's health/ | 133489 |
| 24 | school attendance/ or absenteeism/ | 9595 |
| 25 | health care access/ | 31015 |
| 26 | (predictor* or "risk factor*" or determinant* or "sexual behaviour" or "multiple sex partner*" or "sell* sex*" or extramarital* or "transactional sex" or prostitut* or "sex work" or condom* or "unsafe sex" or "unprotected sex" or knowledge or "social influenc*" or "peer influenc*" or "social norm" or cultur* or sociocultural* or socioeconomic* or "social environmental*" or socioenvironment* or Stigma or discriminat* or "psychological impact" or "social impact" or "psychosocial impact" or stress or distress or depression).tw,kw. | 4647253 |
| 27 | or/10-26 | 5322037 |
| 28 | family/ | 81842 |
| 29 | (family* or families).tw,kw. | 1070817 |
| 30 | 28 or 29 | 1100542 |
| 31 | developing country/ | 79007 |
| 32 | ((Developing or Less developed or low resource* or disadvantaged or resource limited or poor or low* or middle income*) adj (countr* or region* or nation? or area*)).tw,kw. | 89188 |
| 33 | 31 or 32 | 144881 |
| 34 | 4 and 9 and 27 and 30 and 33 | 841 |
| 35 | limit 34 to yr="2004 - 2021" | 183 |

Database(s): **CINAHL**
Search Strategy:

| **#** | **Searches** | **Results** |
| --- | --- | --- |
| S1 | (MH "HIV-Infected Patients") OR (MH "Human Immunodeficiency Virus") OR (MH "HIV-1") | 10,938 |
| S2 | TI ( (HIV* or "Human immunodeficiency virus" or AIDS) ) OR AB ( (HIV* or "Human immunodeficiency virus" or AIDS) ) | 129,961 |
| S3 | (MH "Acquired Immunodeficiency Syndrome") OR (MH "AIDS Patients") | 19,582 |
| S4 | S1 OR S2 OR S3 | 135,254 |
| S5 | (MH "Female") | 2,132,233 |
| S6 | (MH "Married Women") OR (MH "Single Women") OR (MH "Women") | 22,887 |
| S7 | (MH "Mothers") OR (MH "Expectant Mothers") OR (MH "Adolescent Mothers") OR (MH "Surrogate Mothers") | 46,888 |
| S8 | TI ( (Wives or Wife or Mothers or female* or girl* or wom?n) ) OR AB ( (Wives or Wife or Mothers or female* or girl* or wom?n) ) | 749,596 |
| S9 | S5 OR S6 OR S7 OR S8 | 2,303,417 |
| S10 | (MH "Sexual Abuse") | 9,465 |
| S11 | (MH "Unsafe Sex") | 3,481 |
| S12 | (MH "Sexual Harassment") | 2,227 |
| S13 | (MH "Bisexuality") | 1,492 |
| S14 | (MH "Heterosexuality") | 2,073 |
| S15 | (MH "Risk Factors") | 196,278 |
| S16 | (MH "Sexual Partners") | 9,882 |
| S17 | (MH "Prostitution") OR (MH "Unsafe Sex") | 3,481 |
| S18 | (MH "Condoms") | 8,161 |
| S19 | (MH "Socioeconomic Factors") OR (MH "Illiteracy") OR (MH "Poverty") OR (MH "Educational Status") OR (MH "Employment") OR (MH "Unemployment") OR (MH "Social Norms") OR (MH "Social Networks") | 193,503 |
| S20 | (MH "Stigma") | 18,823 |
| S21 | (MH "Discrimination") | 12,231 |
| S22 | (MH "Psychosocial Aspects of Illness") OR (MH "Morals") OR (MH "Social Isolation") | 22,863 |
| S23 | (MH "Stress") OR (MH "Stress, Psychological") | 66,095 |
| S24 | (MH "Depression") | 120,367 |
| S25 | (MH "Income") | 22,095 |
| S26 | (MH "Adolescent Nutrition") OR (MH "Child Nutrition") OR (MH "Nutritional Status") OR (MH "Infant Nutrition") | 27,865 |
| S27 | (MH "Adolescent Health") OR (MH "Child Health") OR (MH "Family Health") OR (MH "Health Status") OR (MH "Mental Health") OR (MH "Wellness") | 135,966 |
| S28 | (MH "Health Services Accessibility") | 97,272 |
| S29 | (MH "Absenteeism") | 5,033 |
| S30 | (MH "Psychological Well-Being") | 30,301 |
| S31 | TI ( (predictor* or "risk factor*" or determinant* or "sexual behaviour" or "multiple sex partner*" or "sell* sex*" or extramarital* or "transactional sex" or prostitut* or "sex work" or condom* or "unsafe sex" or "unprotected sex" or knowledge or "social influenc*" or "peer influenc*" or "social norm" or cultur* or sociocultural* or socioeconomic* or "social environmental*" or socioenvironment* or Stigma or discriminat* or "psychological impact" or "social impact" or education or "school attendance" or "withdraw* from school" or "psychosocial impact" or stress or distress or depression or employment or "loss of job" or Income or "nutrition security" or "food insecurity" or health or "physical health" or wellbeing or "Healthcare accessibility*" or Absenteeism or religio* or Consequence*) ) OR AB ( (predictor* or "risk factor*" or determinant* or "sexual behaviour" or "multiple sex partner*" or "sell* sex*" or extramarital* or "transactional sex" or prostitut* or "sex work" or condom* or "unsafe sex" or "unprotected sex" or knowledge or "social influenc*" or "peer influenc*" or "social norm" or cultur* or sociocultural* or socioeconomic* or "social environmental*" or socioenvironment* or Stigma or discriminat* or "psychological impact" or "social impact" or "psychosocial impact" or stress or distress or depression) ) | 1,549,822 |
| S32 | S10 OR S11 OR S12 OR S13 OR S14 OR S15 OR S16 OR S17 OR S18 OR S19 OR S20 OR S21 OR S22 OR S23 OR S24 OR S25 OR S26 OR S27 OR S28 OR S29 OR S30 OR S31 | 1,936,189 |
| S33 | (MH "Family") | 44,723 |
| S34 | TI ( (family* or families) ) OR AB ( (family* or families) ) | 272,246 |
| S35 | S33 OR S34 | 287,870 |
| S36 | (MH "Developing Countries") OR (MH "Indian Ocean Islands") OR (MH "Low and Middle Income Countries") OR (MH "Asia") | 28,756 |
| S37 | TI ( ((Developing or Less developed or low resource* or disadvantaged or resource limited or poor or low* income*) N (countr* or region* or nation? or area*)) ) OR AB ( ((Developing or Less developed or low resource* or disadvantaged or resource limited or poor or low* or middle income*) N (countr* or region* or nation? or area*)) ) | 97 |
| S38 | S36 OR S37 | 28,818 |
| S39 | S4 AND S9 AND S32 AND S35 AND S38 (Published Date: 20040101-20221231) | 47 |

Database(s): **PsycINFO**
Search Strategy:

| **#** | **Searches** | **Results** |
| --- | --- | --- |
| 1 | hiv/ or "aids (attitudes toward)"/ or aids prevention/ or *HIV Testing/ | 43172 |
| 2 | (HIV* or "Human immunodeficiency virus" or AIDS).ti,ab,id. | 75092 |
| 3 | 1 or 2 | 75333 |
| 4 | wives/ or human females/ or spouses/ | 109509 |
| 5 | exp EXPECTANT MOTHERS/ or exp UNWED MOTHERS/ or exp ADOLESCENT MOTHERS/ or exp SINGLE MOTHERS/ or exp MOTHERS/ | 44768 |
| 6 | (Wives or Wife or Mothers or female* or girl* or wom?n).ti,ab,id. | 746674 |
| 7 | 4 or 5 or 6 | 759753 |
| 8 | exp Condoms/ or exp Sexual Risk Taking/ or exp Sexual Partners/ | 15923 |
| 9 | exp Sexual Partners/ | 5115 |
| 10 | risk factors/ or psychosocial factors/ | 125029 |
| 11 | socioeconomic status/ or family socioeconomic level/ or income level/ or lower class/ or social class/ or disadvantaged/ or economic security/ or "income (economic)"/ or poverty/ | 62724 |
| 12 | exp Sociocultural Factors/ | 126837 |
| 13 | exp Emotional Trauma/ or exp Stress/ or exp Distress/ or exp Psychological Stress/ or exp Well Being/ | 211167 |
| 14 | major depression/ | 137841 |
| 15 | exp DISCRIMINATION/ or exp SOCIAL DISCRIMINATION/ | 52937 |
| 16 | stigma/ | 15171 |
| 17 | (predictor* or "risk factor*" or determinant* or "sexual behaviour" or condom* or "unsafe sex" or "unprotected sex" or "transactional sex" or prostitute* or "social influence" or "social norm" or cultur* or "sociocultural factor*" or socioeconomic* or "social environmental*" or socioenvironment* or Stigma or discrimination or "Psychological impact" or "Social impact" or Education or "Psychosocial impact" or stress or distress or depression).ti,ab,id. | 1501319 |
| 18 | or/8-17 | 1662981 |
| 19 | family conflict/ or mother-child relations/ or parenting/ | 37831 |
| 20 | *family/ | 33154 |
| 21 | (family* or families).ti,ab,id. | 407965 |
| 22 | 19 or 20 or 21 | 432552 |
| 23 | 3 and 7 and 18 and 22 | 1799 |
| 24 | exp Developing Countries/ | 6015 |
| 25 | ((Developing or Less developed or low resource* or disadvantaged or resource limited or poor or low* or middle income*) adj (countr* or region* or nation? or area*)).ti,ab,id. | 17343 |
| 26 | 24 or 25 | 18829 |
| 27 | 23 and 26 | 53 |

Database(s): **Ovid MEDLINE(R)**
Search Strategy:

| **#** | **Searches** | **Results** |
| --- | --- | --- |
| 1 | exp HIV/ or HIV-1/ or HIV-2/ or HIV INFECTIONS/ | 260283 |
| 2 | Acquired Immunodeficiency Syndrome/ | 77403 |
| 3 | (HIV* or "Human immunodeficiency virus" or AIDS).tw,kf. | 439882 |
| 4 | or/1-3 | 474973 |
| 5 | mothers/ or surrogate mothers/ | 52441 |
| 6 | Female/ | 9391282 |
| 7 | Women/ | 15138 |
| 8 | (Wives or Wife or Mothers or female* or girl* or wom?n).tw,kf. | 2463234 |
| 9 | or/5-8 | 9812155 |
| 10 | sexual behavior/ or extramarital relations/ or hiv serosorting/ or sex work/ or safe sex/ or sexual harassment/ or bisexuality/ or heterosexuality/ or homosexuality/ or transsexualism/ or unsafe sex/ | 91745 |
| 11 | Condoms/ | 10909 |
| 12 | Sexual Partners/ | 19299 |
| 13 | culture/ or social environment/ or social isolation/ or social norms/ or socioeconomic factors/ or poverty/ or poverty areas/ or social class/ | 321694 |
| 14 | SOCIAL STIGMA/ | 11233 |
| 15 | "Discrimination (Psychology)"/ | 19915 |
| 16 | STRESS, PSYCHOLOGICAL/ | 130419 |
| 17 | DEPRESSION/ | 139209 |
| 18 | (predictor* or "risk factor*" or determinant* or "sexual behaviour" or "multiple sex partner*" or extramarital* or "sell* sex*" or "transactional sex" or prostitut* or "sex work" or condom* or "unsafe sex" or "unprotected sex" or knowledge or "social influenc*" or "peer influenc*" or "social norm" or cultur* or socioeconomic* or "social environmental*" or socioenvironment* or Stigma or discriminat* or "Psychological impact" or "Social impact" or education or "school attendance" or "withdraw* from school" or Stress or "Psychosocial impact" or stress or depression or distress).tw,kf. | 5029349 |
| 19 | or/10-18 | 5299336 |
| 20 | family/ or family relations/ | 92433 |
| 21 | (family* or families).tw,kf. | 1073772 |
| 22 | 20 or 21 | 1106485 |
| 23 | Developing Countries/ | 79007 |
| 24 | ((Developing or Less developed or low resource* or disadvantaged or resource limited or poor or low* or middle income*) adj (countr* or region* or nation? or area*)).tw,kf. | 137093 |
| 25 | 23 or 24 | 169922 |
| 26 | 4 and 9 and 19 and 22 and 25 | 1254 |
| 27 | limit 26 to (english language and yr="2004 - 2021") | 226 |

**Database: SCOPUS**

Search strategy:

TITLE-ABS-KEY ((hiv* OR "Human immunodeficiency virus" OR aids)) AND TITLE-ABS-KEY ((wives OR wife OR mothers OR female* OR girl* OR women OR woman)) AND TITLE-ABS-KEY ((predictor* OR "risk factor*" OR determinant* OR "sexual behaviour" OR "multiple sex partner*" OR extramarital* OR "sell* sex*" OR "transactional sex" OR prostitut* OR "sex work" OR condom* OR "unsafe sex" OR "unprotected sex" OR knowledge OR "social influenc*" OR "peer influenc*" OR "social norm*" OR cultur* OR sociocultural* OR socioeconomic* OR "social environmental*" OR socioenvironment* OR stigma OR discriminat* OR "psychological impact" OR "social impact" OR stress OR distress OR depression OR "psychosocial impact")) AND TITLE-ABS-KEY ((family* OR families)) AND TITLE-ABS-KEY (((developing OR "Less developed" OR "low resource*" OR disadvantaged OR "resource limited" OR poor OR "low* OR middle income*") W/0 (countr* OR region* OR nation? OR area*))) AND (LIMIT-TO (PUBYEAR, 2021) OR LIMIT-TO (PUBYEAR, 2004)) AND (LIMIT-TO (LANGUAGE, "English"))

Results: 30

**Database(s): ProQuest**

Search strategy:

noft((HIV* OR "Human immunodeficiency virus" OR AIDS) AND (Wives OR Wife OR Mothers OR female* OR girl* OR wom?n) AND (predictor* OR "risk factor*" OR determinant* OR "sexual behaviour" OR "multiple sex partner*" OR "sell* sex*" OR "transactional sex" OR prostitut* OR "sex work" OR condom* OR "unsafe sex" OR "unprotected sex" OR knowledge OR "social influenc*" OR "peer influenc*" OR "social norm" OR cultur* OR sociocultural* OR socioeconomic* OR "social environmental*" OR socioenvironment* OR Stigma OR discriminat* OR "Psychological impact" OR "Social impact" OR "Psychosocial impact" OR distress OR stress OR Depression) AND (family* OR families) AND ((Developing OR "Less developed" OR "low resource*" OR disadvantaged OR "resource limited" OR poor OR "low* OR middle income*") NEAR/0 (countr* OR region* OR nation? OR area*))) AND PEER(yes)

Results: 272

**Appendix 2: JBI Critical Appraisal instruments**

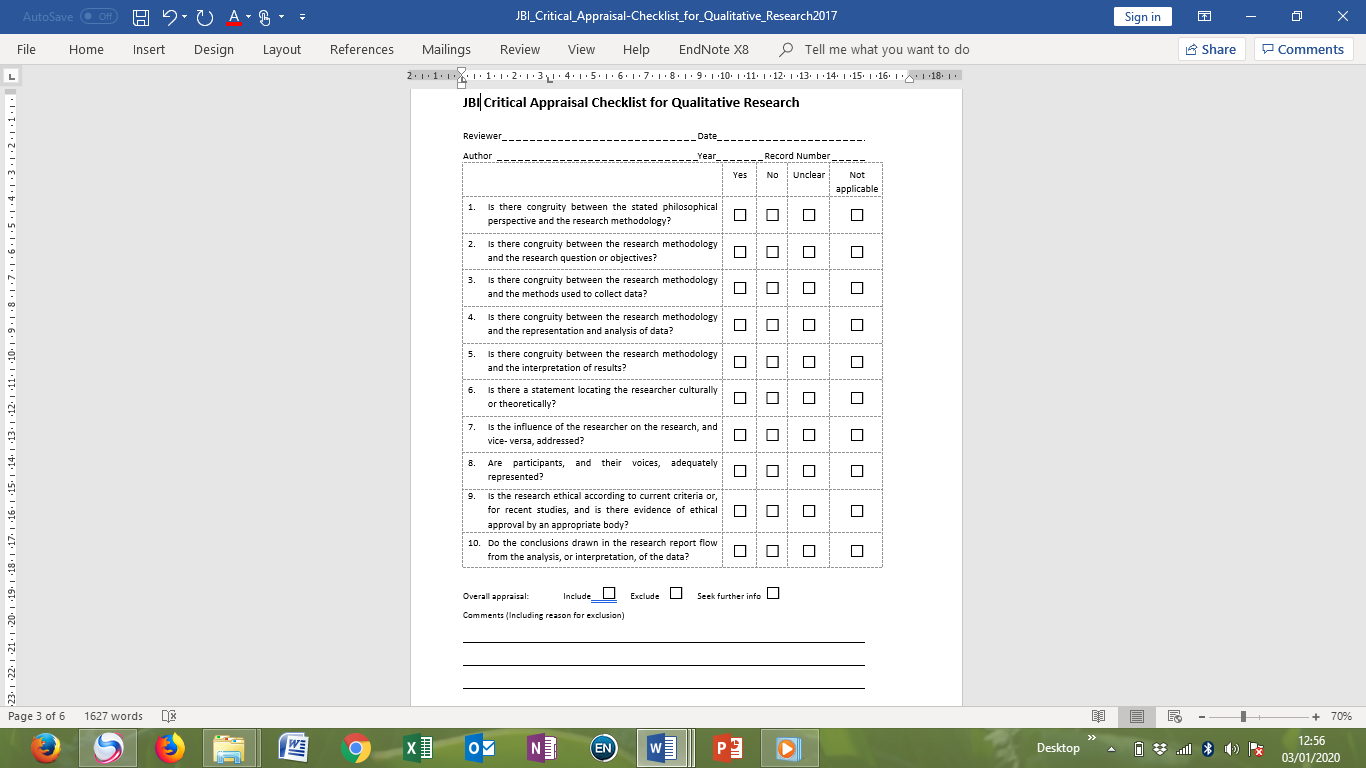

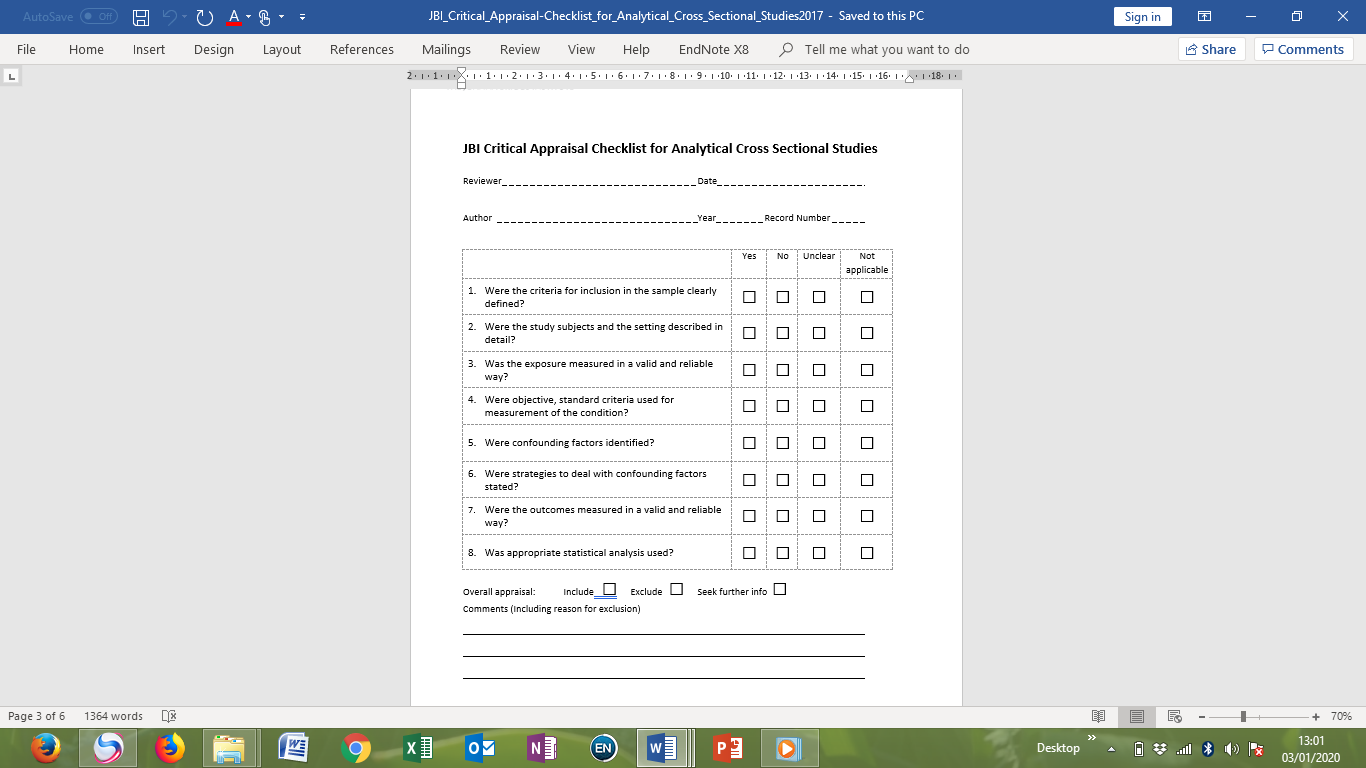

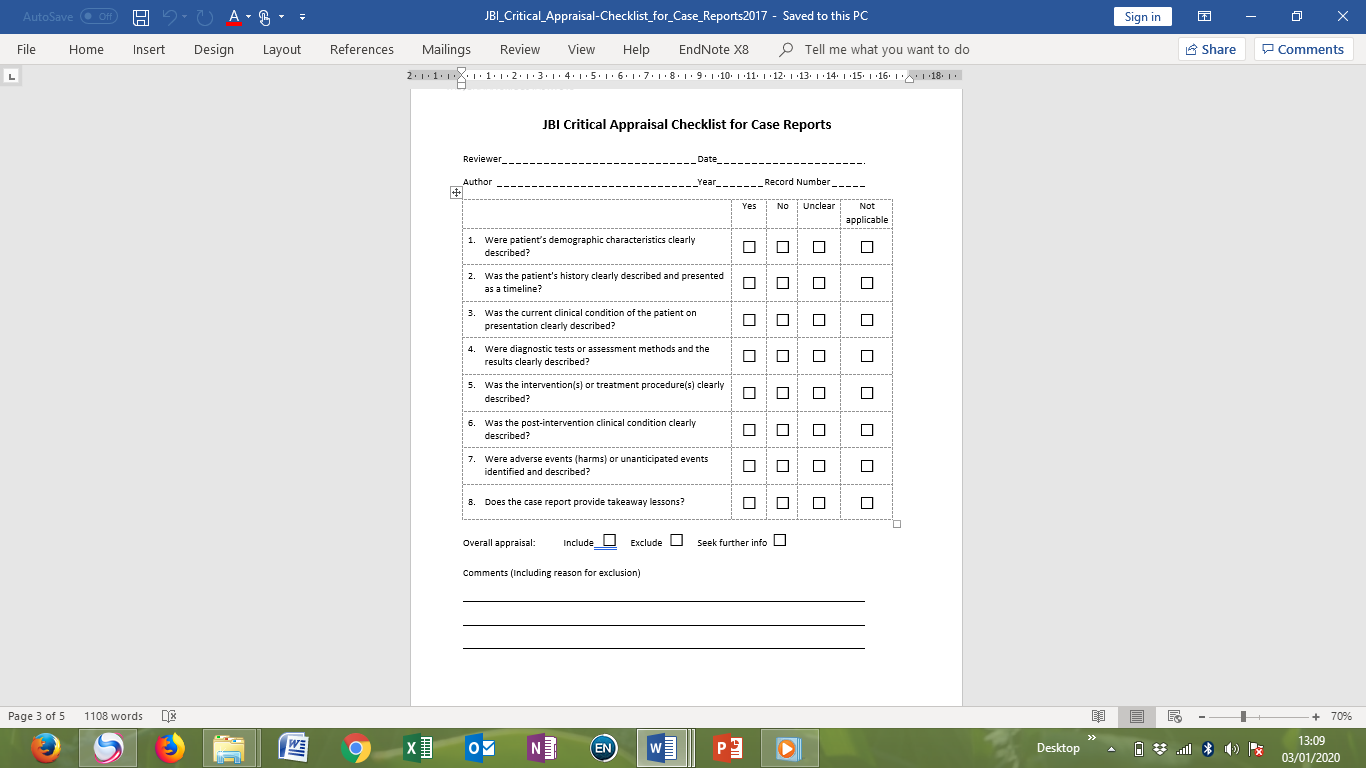

**Appendix 3: Assessment of methodological quality (n=17)**

| **Authors** | **Q1** | **Q2** | **Q3** | **Q4** | **Q5** | **Q6** | **Q7** | **Q8** | **Q9** | **Q10** | **%** |
| --- | --- | --- | --- | --- | --- | --- | --- | --- | --- | --- | --- |
| Azhar | Y | Y | Y | Y | Y | Y | U | Y | Y | Y | 100 |
| Chi et al | Y | Y | Y | Y | Y | N | Y | Y | Y | Y | 90 |
| de Souza | Y | Y | Y | Y | Y | Y | Y | Y | N | Y | 90 |
| Fauk et al | Y | Y | Y | Y | Y | Y | Y | Y | Y | Y | 100 |
| Halli et al | Y | Y | Y | Y | Y | Y | Y | Y |  |  | 100 |
| Halimatusa’diah | Y | Y | Y | Y | Y | Y | Y | Y | N | Y | 90 |
| Ismail et al | Y | Y | Y | Y | Y | Y | Y | Y | Y | Y | 100 |
| Liamputtong et al | Y | Y | Y | Y | Y | Y | Y | Y | Y | Y | 100 |
| Mathew et al | Y | Y | Y | Y | Y | Y | U | Y | Y | Y | 100 |
| Nguyen et al | Y | Y | Y | Y | Y | N | Y | Y | Y | Y | 90 |
| Paxton et al | Y | Y | Y | Y | Y | Y | U | Y |  |  | 100 |
| Qin et al | Y | Y | Y | Y | Y | U | Y | Y |  |  | 100 |
| Srivastava et al | Y | Y | Y | Y | Y | Y | Y | Y | Y | Y | 100 |
| Subramaniyan et al | Y | Y | Y | Y | Y | N | U | Y | Y | Y | 89 |
| Thomas et al | Y | Y | Y | Y | Y | Y | Y | Y | Y | Y | 100 |
| Zang et al | Y | Y | Y | Y | Y | Y | Y | Y |  |  | 100 |
| Yang et al | Y | Y | Y | Y | Y | N | U | Y | Y | Y | 89 |

Q= Question; Y= Yes; N= No; U= Unclear
